# Genomic landscape of Ewing sarcoma: a pooled analysis of 538 cases and clinicopathological correlation

**DOI:** 10.64898/2026.06.29.26356801

**Authors:** Laura Romero-Pérez, Clémence Henon, Andreas Ranft, Juan Díaz-Martin, Florencia Cidre-Aranaz, Uta Dirksen, Enrique de Álava, Thomas G. P. Grünewald

## Abstract

**Background:** Ewing sarcoma (EwS) is a highly aggressive bone and soft tissue cancer mainly affecting children, adolescents, and young adults. The rarity of the disease, relatively small cohort sizes of prior studies, and overall low mutational burden of EwS have limited the ability to establish robust correlations of genomic findings and clinicopathological factors.

**Methods:** To overcome these limitations, we integrated genomic and clinical data from the seven major sequencing studies encompassing 538 EwS patients. Mutational profiles (SNV, indels and CNVs), and their correlation with clinicopathological features in the aggregated cohort were systematically analyzed to provide an integrated view of the EwS genomic landscape.

**Results:** This study compiles the largest EwS genomic dataset reported to date. In the aggregated cohort (*n*=538) bone tumors were more common (65.4%) than soft-tissue tumors (34.6%), the latter being more frequent in older male patients and associated with poorer outcomes. EWSR1::FLI1 was the most prevalent fusion (87.2%). No major clinicopathological differences were identified between fusion types. The mutational landscape was dominated by *STAG2* (15.6%) and *TP53* (7.1%) alterations, associated with younger or with older age at diagnosis and poor survival, respectively. Strikingly, the coexistence of *STAG2* and *TP53* mutations, although rare (*n*=12), was associated with lethal outcome in all cases. *CDKN2A* loss (9.1%) was associated with older age, poor survival, and linked to a higher frequency of *TP53*-mutations in soft tissue EwS. Among frequent CNVs, gain of chr1q (25.2%) and loss of chr16q (21.9%) were per se frequently associated with fatal outcome and their co-occurrence further increased the risk of lethality.

**Conclusions:** We delineate recurrent genomic alterations with important clinicopathological associations, including a uniformly lethal *STAG2/TP53* co-mutation and CNV signatures marking aggressive disease. This comprehensive pooled analysis of EwS genomic studies provides a foundation for refined biological risk-stratification.

## INTRODUCTION

Ewing sarcoma (EwS) is a highly aggressive bone or soft tissue cancer mostly affecting children, adolescents, and young adults^1^. Under modern multimodal and interval-compressed treatment regimens, the survival rates for patients with localized disease reach ∼85%. However, the survival rates for patients presenting with metastatic disease at diagnosis remain unacceptably low (<30%)^1^. EwS is genetically well characterized. It is defined by the presence of recurrent translocations involving members of the *FET* (*FUS*, *EWSR1*, *TAF15*) gene family (in most cases *EWSR1*), and a member of the ETS family of transcription factors (in ∼85% of cases *FLI1*, in ∼10% *ERG*, and each in <1% *ETV1*, *ETV4*, or *FEV*)^1^.

During the last decade, several sequencing and genomic efforts have been undertaken to further illuminate the genetic landscape of EwS. At the genomic level, recurrent gains have been described in chromosome (chr) 8, 12, 1q and 20, which may be accompanied by genomic losses such as losses of 16q^1,2^. In addition, multiple older genomic studies comprising discovery sets of 177 (ref.^3^ and ref.^4^), 132 (ref.^5^), 92 (ref.^6^), 65 (ref.^7^) 52 (ref.^8^), and 20 (ref.^9^) patients, and other more recent studies comprising 351 (ref.^10^), 196 (ref.^11^; some of them included in ref.^3^ and ref.^4^) and 23 (ref.^12^) patients, identified recurrent mutations in the *STAG2 Cohesin Complex Component* gene (*STAG2*) in 7.6 –21.5% of cases and in the *Tumor Protein P53* gene (*TP53*) in 5–11% of cases^3–11^, which may be associated with a more aggressive phenotype^4–6,10,11,13,14^. Interestingly, beyond *STAG2* and *TP53,* almost all studies identified multiple other recurrent and potentially pathogenic mutations in a diverse set of genes affecting ∼1–4% of cases^3–9,11^. However, these sample sizes remain underpowered by many clinically relevant associations, given low mutational burden of EwS.

In contrast, The Cancer Genome Atlas (TCGA) project providing multidimensional omics data on large cancer cohorts has ignited hundreds of research projects in a wide variety of relatively rare and common cancer entities thereby having a high impact on cancer research in general.

Unfortunately, at present the TCGA sarcoma dataset does not contain any EwS sample (https://portal.gdc.cancer.gov/projects/TCGA-SARC)^15^. Nevertheless, other initiatives aim to fill this gap. For example, the MSK-IMPACT project^3,4,11^, which recently published genomic and clinical information for >2,000 sarcoma samples and matched normal tissues, includes 179 EwS patients used in some studies^3,4^ (https://www.cbioportal.org/study/summary?id=es_dsrct_msk_2023) and it extends to 196 EwS patients very recently processed by another study^11^. Another example is the recent St. Jude Cloud Pecan initiative with curated data from ∼9,000 pediatric cancer samples, of which 165 EwS samples were genomically analyzed (https://www.stjude.cloud/research-domains/pediatric-cancer). However, even this larger cohort remain insufficient to detect associations involving low-frequency genomic events, and especially those that have been classified as variants of uncertain significance having unknown or inconclusive oncogenic effect in 48% of cases^11^.

To overcome these limitations, we performed a pooled analysis of an aggregated series of EwS cases compiling published sequencing data from the seven major studies and repositories at the time we censored the data (*n*=538 patients). Using this harmonized dataset we confirmed known clinicopathological associations of recurrent *STAG2* and *TP53* mutations as well as chromosomal gains and losses but also uncovered hitherto unknown clinicopathological associations, such as those derived from the concomitance of *STAG2*, *CDKN2A* and *TP53* alterations, those associated with the presence of *ERF* or *EZH2* mutations, or those observed in presence of the combination of *STAG2*, *TP53* or *CDKN2A* alterations with gains of chr1q, chr8, chr12, chr20 or losses of chr16q.. Moreover, our study provides the largest source of genomic information on EwS available to the research community.

## RESULTS

### Aggregated data from published sequencing studies provide a comprehensive view on the genomic landscape of EwS

To obtain a global and more precise view of mutational events in EwS, we aggregated existing published datasets from the most pertinent sequencing studies providing molecular and clinical information^3–9^. This aggregated cohort–henceforth designated ‘study cohort’–comprised mutational data from 538 EwS patients with detailed clinicopathological characteristics for most of them (*n*=315–508, with respect to the given clinicopathological/genomic feature) (**Fig. 1**). The clinicopathological characteristics were comparable with those of previous clinical/epidemiological studies^3–9^ **(Table 1)**. Given the heterogeneity across platforms (WGS, WES, SNP array, targeted sequencing), genomic variables were standardized into binary categories (e.g., mutated/not mutated; gain/loss).

**Figure 1.**
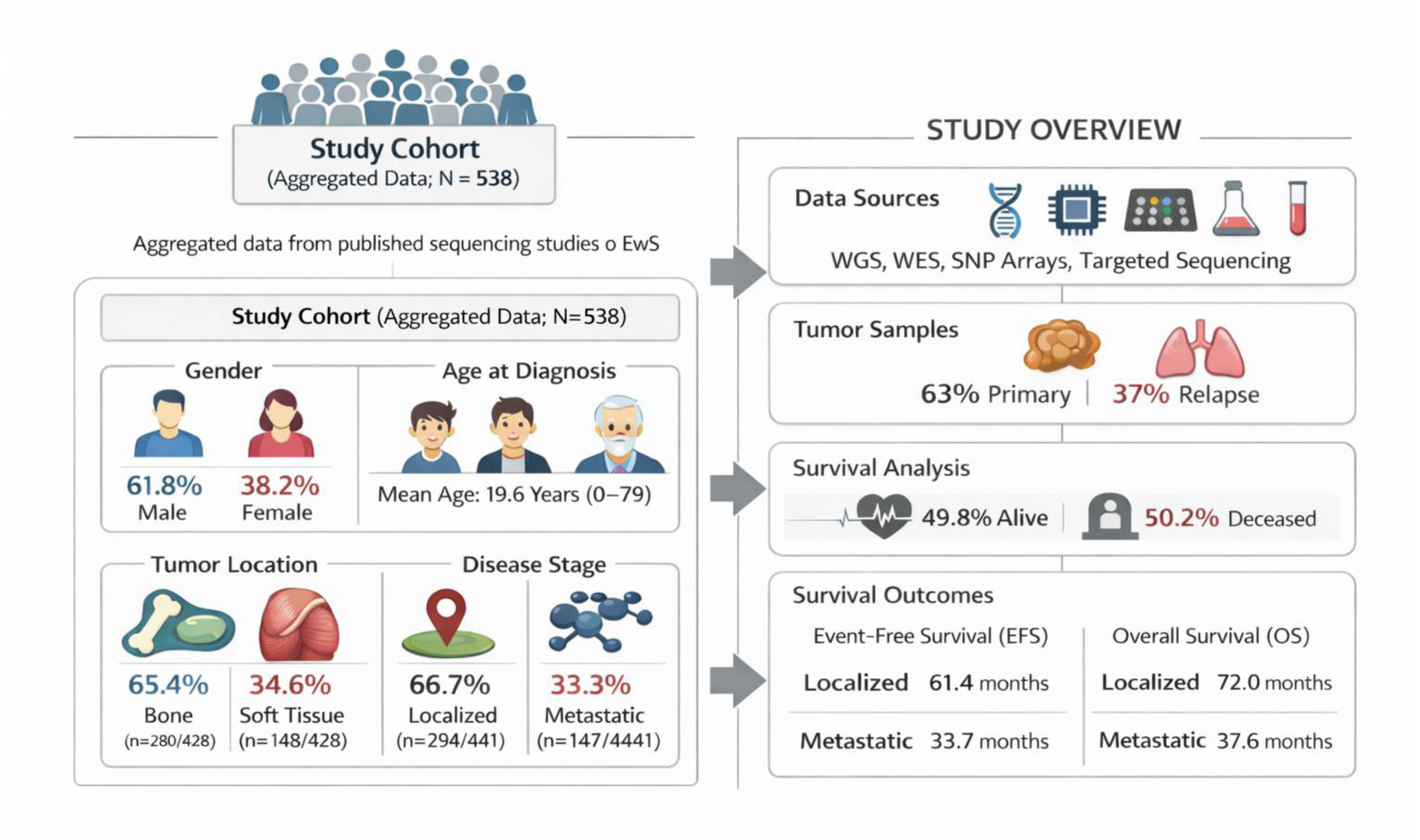
Study cohort overview. Summary of the main representative clinical characteristics of the study cohort.

**Table 1.**
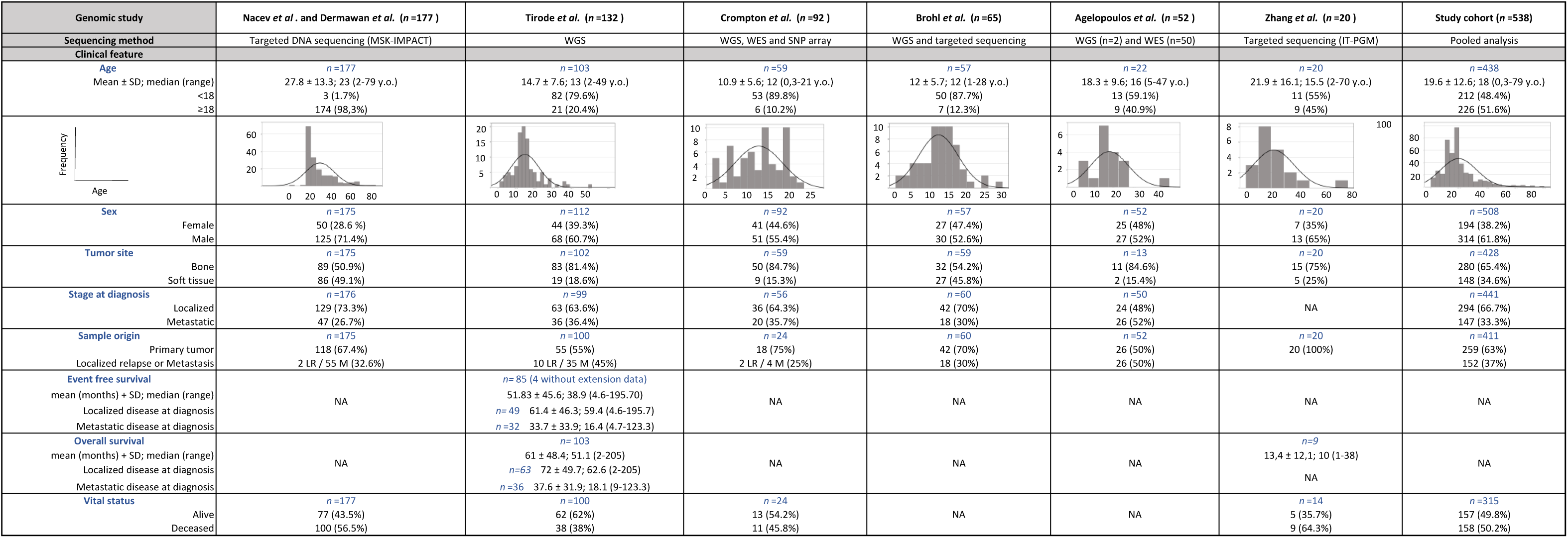
Clinicopathological characteristics and composition of the different studies included in the study cohort. In most cases, the vital status was not accompanied by average follow-up, so these data should be interpreted with caution. NA: Not available information.

#### Clinicopathological characteristics of the aggregated study cohort

From the collected clinical information, we observed that 61.8% of cases were male (314/508) and 38.2% female (194/508), an expected ratio (1.62:1) according to literature^16^. The mean age at diagnosis (*n*=438) was 19.6 years (±12.6) ranging from 0 to 79 years, with a marked peak at the median of 18 years. We noted differences in the distribution of ages per study, from those including only or mostly pediatric patients^5–7^, to those including heterogenous^8,9^ or even a vast majority of adult patients^3,4^ (*P*<0.001; **Table 1**). In line with previous reports^17,18^, our analysis revealed a greater proportion of cases arising in soft tissue in older patients (*P*<0.001; **Supp. Fig. 1A**). In this study cohort, soft tissue tumors were more frequent than reported in the literature (∼10–15%)^1^, accounting for 34.6% (148/428) of cases, while 65.4% (280/428) arose on bone.

At time of diagnosis, 66.7% of cases (294/441) had localized disease, while in 33.3% (147/441) presented with metastases. Precise information about the tumor specimen was not always available: of the 411 cases for which information was available, 63% (259/411) were primary tumors and 37% (152/411) were local (*n*=14) or distant (*n*=138) relapses. At last follow-up, 49.8% of patients (157/315) were alive, while 50.2% died due to EwS (158/315). However, event-free survival (EFS) and overall survival (OS) data were only available for 85 (ref.^5^) or 103 (ref.^5^) patients, respectively. One study reported OS time exclusively for deceased patients^9^, while two other datasets^3,4^ contained inconsistent or inconclusive survival annotation in public repositories. Therefore, these three studies were excluded from the EFS/OS analysis. Among the evaluable cases (ref.^5^), both EFS and OS were, as expected, longer in patients diagnosed with localized disease compared with those presenting with metastatic disease (EFS: 61.4±46.3 *versus* 33.7±33.9 months; HR: 2.15; 95% CI: 1.16−3.98; *P*=0.015; and OS: 72±49.7 *versus* 37.6±18.1 months; HR: 2.76; 95% CI: 1.46−5.2; *P*=0.002, respectively, **Supp. Figs. 1B-C**).

#### Soft tissue EwS are associated with older male patients and poor prognosis

Analyses of associations among clinical parameters revealed a higher frequency of tumors arising in soft tissue in older patients (22.6±14.2 versus 18±11.5; *P*<0.001. **Supp. Fig. 1A**) and in the male population (72%; 105/146) compared with females (28%; 41/146; *P*<0.01). In addition, the proportion of patients dying of EwS was higher among patients with extraosseous EwS as compared to patients with bone EwS [58.5% (65/111) *versus* 43.7% (88/201), respectively; *P*<0.05], as was previously published^19,20^. This did not translate into strong sex-specific differences in survival (*P*=0.12).

#### FET::ETS fusion types are not associated with specific clinicopathological features

Previous studies have reported conflicting results regarding the potential association between the FET::ETS fusion type and outcome parameters, including chromoplexy or the induction of some differences in the transcriptional profile^21–23^. Since our study cohort contained precise information on the fusion type for 374 of 538 samples, we sought to investigate potential associations with clinicopathological features. In this large study cohort, *EWSR1::FLI1* was detected in 87.2% (326/374) of cases, *EWSR1::ERG* in 10.9% (41/374), *EWSR1::FEV* in 0.8% (3/374), *FUS::FEV* in 0.5% (2/374), and *EWSR1::ETV1* and *EWSR1::ETV4* in 0.3% each (1/374).

In line with previously reported findings, EwS tumors harboring either *EWSR1::FLI1* or *EWSR1::ERG* fusions presented comparable clinical phenotypes and outcomes^1^ **(Supp. Table 1)**. Patients with tumors bearing either fusion had a similar age at diagnosis (mean of 20.5±12.6 and 22.4±13.6 years old, respectively; *P*=.0.42) and proportion of sexes [64.8 (205/316) and 63.4% (26/41) of males, and 35.1% (111/316) and 36.6% (15/41) of females, respectively. *P*=0.76]. We found no relevant differences when analyzing either the tissue of origin, the tumor extension at diagnosis, or the proportion of deceased patients. Although the low number of cases with alternative *FET::ETS* fusions [*EWSR1::FEV* (*n*=3); *FUS::FEV* (*n*=2); *EWSR1::ETV1* (*n*=1); *EWSR1::ETV4* (*n*=1)] precluded statistical analysis for these cases, our pooled analysis further supports the notion that the major fusion types have no prognostic value.

#### The mutational profile of EwS is dominated by STAG2 and TP53 mutations

*STAG2* was the most frequently mutated gene (15.6%; 84/538), followed by *Titin (TTN*, 11.7%; 38/324) and *TP53* (7.1%; 38/538) (**Fig. 2**). *TTN* is a large gene encoding one of the largest human proteins (36,800 aa), found to be recurrently mutated in most cancer genome studies^24^ and in cancer-free control groups^25^, and no relevant role in EwS oncogenesis has been reported. Therefore, we excluded *TTN* from further analyses.

**Figure 2.**
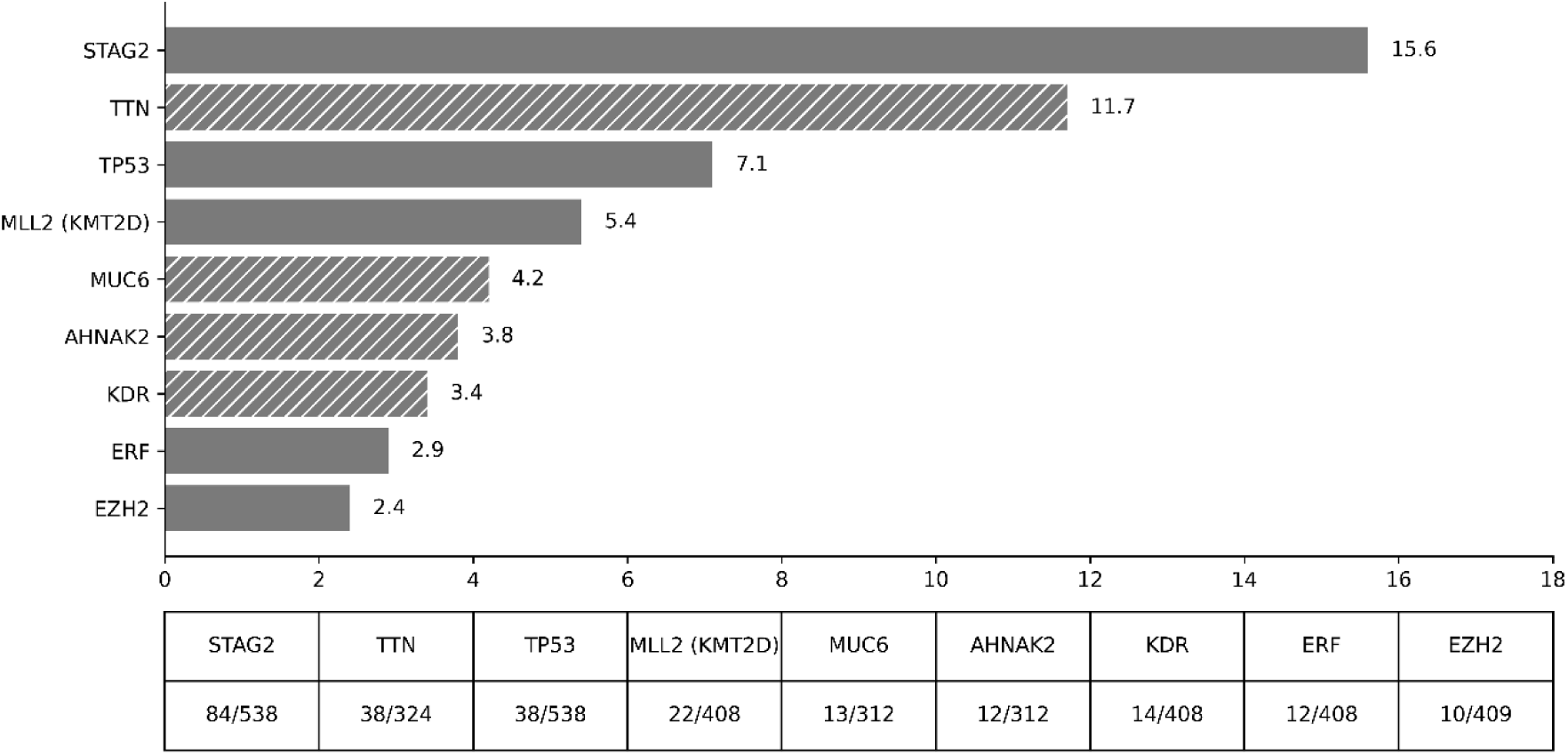
Most frequently mutated genes in the study cohort. Hatched bars denote genes excluded from clinicopathological correlation analyses for predefined reasons. The table summarizes mutation frequency as the number of affected cases over the total number of evaluable samples.

In the case of *Mucin-6* (*MUC6*) or *AHNAK Nucleoprotein 2* (*AHNAK2*), found to be mutated in 4.2% (13/312) and 3.8% (12/312) of cases, respectively, most of patients (12/13 and 12/12, respectively) derived from the same study^6^, which reported that both mutated genes were detected by WES, but these variants were not confirmed by complementary sequencing (WGS) or SNP array profiling, wherefore they have not been considered for further analyses. Instead, *Lysine methyltransferase 2D (MLL2,* now *KMT2D)* variants, present in 5.4% of cases in our study cohort (22/408; 16 of which were also found in ref^6^), were validated^6^, being all different mutations. Thus, we decided to keep *MLL2* in our study. MLL2 (KMT2D) acts as a transcriptional co-activator, crucial for enhancer activation and gene expression during development^26^.

For several reasons, we decided not to consider *KDR*, which encodes the Kinase Insert Domain Receptor, involved in angiogenesis: Mutations in the *KDR* gene were almost exclusively detected in the study of Zhang et al.^9^ (13/14 affected patients), being present in 2.8% (14/503) of our study cohort. Since these cases comprised mainly the same missense variants [85.7% of cases (12/14; Q472H, c.1416A>T, confirmed by Sanger-sequencing)], all derived from the same center (Pekin University Third Hospital, China)^9^, we evaluated whether this observation may relate to a specific genetic/ethnic background. The NCBI dbSNP database reports this variant as a germline polymorphism (rs1870377) with an alternative allele frequency of 0.4730 in the Asian population (0.4473 in East Asian and 0.578 in other Asian), which is much higher than in the European population (0.239558). Despite this missense variant (Q472H; c.1416A>T) is reported in COSMIC (COSV55758724) to be associated with other malignancies, such as melanoma^27^, nasopharyngeal carcinoma^28^, diffuse large B cell lymphoma^29^, or embryonal rhabdomyosarcoma^30^, it is predicted to be likely benign (https://franklin.genoox.com and PolyPhen-2 v2.2.3r406 http://genetics.bwh.harvard.edu/pph2/index.shtml, score=0.003).

By contrast, alterations in *STAG2*, *TP53*, *ERF*, or *EZH2* were not restricted to an individual study, but consistently detected in several reports. *ERF* gene (12/408; 2.9%), encodes the ETS2 Repressor Factor, a strong repressor of ETS-domain proteins, regulating stem cell differentiation, cell fate, and gene expression. *EZH2* (10/409; 2.4%) encodes the Enhancer of Zeste 2 Polycomb Repressive Complex 2 (PRC2) Subunit, a histone methyltransferase of the PRC2 complex, which plays a critical role in epigenetic silencing. Both genes have been previously described as mutated in the context of EwS^5,6,31,32^.

Notably, 35.8% (34/95) of the variants observed in *STAG2* were likely deleterious [i.e. 17.9% (17/95) frameshift deletions/insertions, 9.5% (19/95) splice variants, 8.4% (8/95) missense] and 31.6% were nonsense variants (30/95, with R216X and R259X being the most frequent ones). The remaining *STAG2* variants were detected in the 3’-UTR (15.8%; 15/95), in-frame deletions (2%; 2/95), or not specified (14.7 %; 14/95). In contrast, most of the detected variants in *TP53* were likely deleterious (71%; 32/45) and nonsense variants (4.44%; 2/45), although 24.4% (11/45) were not specified. Two cases were annotated as harboring *TP53*-deep deletions, and we classified these as *TP53* loss-of-function alterations.

#### Clinicopathological implications of the most frequently mutated genes

##### STAG2 mutations are associated with younger age and a trend to appear with metastatic disease at diagnosis

To evaluate the potential clinical implications of *STAG2* and other mutated genes, we correlated available clinicopathological characteristics such as sex (*n*=508), age at diagnosis (*n*=438), disease stage at diagnosis (localized *versus* metastatic; *n*=441), tumor site (bone *versus* soft tissue; *n*=428) or patient status (alive *versus* dead; *n*=315), as well as categorizing features such as the oncogenic fusion type, with the respective mutational data of the ‘study cohort’.

Several individual reports have indicated an association between *STAG2* mutations and metastasis^5,6,10,11,13,14^. According to the collected data, *STAG2* was mutated in 15.6% of EwS samples (84/538; **Fig. 2**). The presence of *STAG2* mutations tends to be associated with metastatic disease (*P*=0.08. **Table 2a**). Indeed, 17.3% (26/150) of metastatic cases at diagnosis harbored *STAG2* mutations, whereas only 11.3% (33/291) of localized tumors did (OR=1.64, 95% CI: 0.94−2.86, *P*=0.08). In addition, patients harboring *STAG2* mutations were, on average, younger at diagnosis than those with non-*STAG2* mutated tumors (15.8±9.5 *versus* 20.2±12.9 years old; *P*<0.05). These results were confirmed by a logistic regression analysis: increasing age was associated with lower odds of *STAG2* mutation (OR 0.96, 95% CI:0.93–0.99, *P*=0.014). As mentioned above, disease extension at diagnosis showed a trend towards higher odds of *STAG2* mutation compared with localized disease (**Table 2a**). When we evaluated the clinicopathological implications segregating by disease stage at diagnosis (**Tables 2b, c**), we observed that the age at diagnosis of *STAG2* mutated EwS cases was lower only in patients with localized disease (mean age of 13.8±10.1; OR 0.94; 95% CI:0.90–0.98; *P*=0.009; **Table 2b**).

**Table 2.**
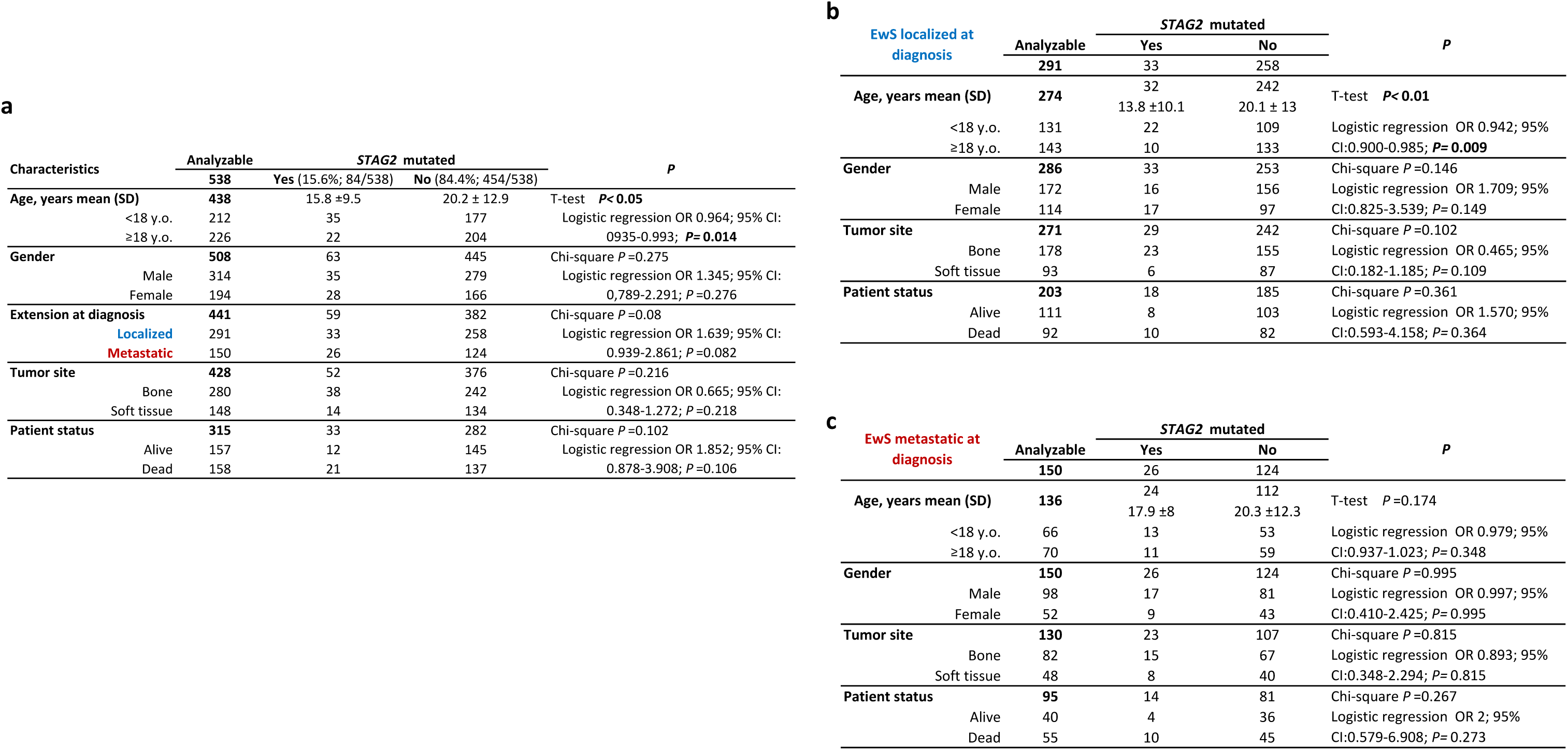
Clinicopathological implications of the presence of *STAG2* mutations: **a)** Global study cohort; **b)** Localized cases. **c)** Metastatic cases.

**a**
| Characteristics | Analyzable | STAG2 mutated |  | P |
| --- | --- | --- | --- | --- |
|  | 538 | Yes (15.6%; 84/538) | No (84.4%; 454/538) |  |
| Age, years mean (SD) | 438 | 15.8 ± 9.5 | 20.2 ± 12.9 | T-test **P< 0.05** |
| <18 y.o. | 212 | 35 | 177 | Logistic regression OR 0.964; 95% CI: 0.935-0.993; **P= 0.014** |
| ≥18 y.o. | 226 | 22 | 204 |  |
| Gender | 508 | 63 | 445 | Chi-square **P=0.275** |
| Male | 314 | 35 | 279 | Logistic regression OR 1.345; 95% CI: 0.789-2.291; **P=0.276** |
| Female | 194 | 28 | 166 |  |
| Extension at diagnosis | 441 | 59 | 382 | Chi-square **P=0.08** |
| Localized | 291 | 33 | 258 | Logistic regression OR 1.639; 95% CI: 0.939-2.861; **P=0.082** |
| Metastatic | 150 | 26 | 124 |  |
| Tumor site | 428 | 52 | 376 | Chi-square **P=0.216** |
| Bone | 280 | 38 | 242 | Logistic regression OR 0.665; 95% CI: 0.348-1.272; **P=0.218** |
| Soft tissue | 148 | 14 | 134 |  |
| Patient status | 315 | 33 | 282 | Chi-square **P=0.102** |
| Alive | 157 | 12 | 145 | Logistic regression OR 1.852; 95% CI: 0.878-3.908; **P=0.106** |
| Dead | 158 | 21 | 137 |  |

**b**
| EwS localized at diagnosis | Analyzable | STAG2 mutated |  | P |
| --- | --- | --- | --- | --- |
|  |  | Yes | No |  |
|  | 291 | 33 | 258 |  |
| Age, years mean (SD) | 274 | 32 | 242 | T-test **P< 0.01** |
|  |  | 13.8 ± 10.1 | 20.1 ± 13 |  |
| <18 y.o. | 131 | 22 | 109 | Logistic regression OR 0.942; 95% CI: 0.900-0.985; **P= 0.009** |
| ≥18 y.o. | 143 | 10 | 133 |  |
| Gender | 286 | 33 | 253 | Chi-square **P=0.146** |
| Male | 172 | 16 | 156 | Logistic regression OR 1.709; 95% CI: 0.825-3.539; **P= 0.149** |
| Female | 114 | 17 | 97 |  |
| Tumor site | 271 | 29 | 242 | Chi-square **P=0.102** |
| Bone | 178 | 23 | 155 | Logistic regression OR 0.465; 95% CI: 0.182-1.185; **P=0.109** |
| Soft tissue | 93 | 6 | 87 |  |
| Patient status | 203 | 18 | 185 | Chi-square **P=0.361** |
| Alive | 111 | 8 | 103 | Logistic regression OR 1.570; 95% CI: 0.593-4.158; **P= 0.364** |
| Dead | 92 | 10 | 82 |  |

| EwS metastatic at diagnosis | Analyzable | STAG2 mutated |  | P |
| --- | --- | --- | --- | --- |
|  |  | Yes | No |  |
|  | 150 | 26 | 124 |  |
| Age, years mean (SD) | 136 | 24 | 112 | T-test **P=0.174** |
|  |  | 17.9 ± 8 | 20.3 ± 12.3 |  |
| <18 y.o. | 66 | 13 | 53 | Logistic regression OR 0.979; 95% CI: 0.937-1.023; **P= 0.348** |
| ≥18 y.o. | 70 | 11 | 59 |  |
| Gender | 150 | 26 | 124 | Chi-square **P=0.995** |
| Male | 98 | 17 | 81 | Logistic regression OR 0.997; 95% CI: 0.410-2.425; **P= 0.995** |
| Female | 52 | 9 | 43 |  |
| Tumor site | 130 | 23 | 107 | Chi-square **P=0.815** |
| Bone | 82 | 15 | 67 | Logistic regression OR 0.893; 95% CI: 0.348-2.294; **P= 0.815** |
| Soft tissue | 48 | 8 | 40 |  |
| Patient status | 95 | 14 | 81 | Chi-square **P=0.267** |
| Alive | 40 | 4 | 36 | Logistic regression OR 2; 95% CI: 0.579-6.908; **P= 0.273** |
| Dead | 55 | 10 | 45 |  |

In our study cohort, we did not observe relevant differences regarding EFS (61.5±14.5 in *STAG2* mutated *versus* 98.8±11 months in *STAG2* wt; HR 1.19; 95% CI:0.55−2.56; *P*=0.659. **Supp. Fig. 2A**), OS (64.9±13.8 in *STAG2* mutated *versus* 104.9±10.7 months in *STAG2* wt; HR 1.33; 95% CI: 0.65−2.73; *P*=0.435. **Supp. Fig. 2B**) or vital status (OR 1.85; *P*=0.106) between *STAG2*-mutated or non-mutated cases (**Fig. 4**). Similarly, when segregating localized *versus* metastatic disease at diagnosis, we found that the presence of *STAG2* mutations not highly increase the risk of relapse (HR 1.28; *P*=0.692 for localized and HR 0.76 and *P*=0.620 for metastatic cases). The presence of *STAG2* mutations was associated with a higher hazard of death in patients with localized disease (HR = 2.14), although this association did not reach statistical significance (*P* = 0.174). No such association was observed in patients with metastatic disease (HR = 0.92, *P* = 0.88). Also, no strong association of *STAG2* mutations and major fusion types were found as 12.6% (41/326) of *EWSR1::FLI1* cases and 19.5% (8/41) of *EWSR1::ERG* cases harbored *STAG2* mutations (*P*=0.22). *STAG2* was found to be mutated in one case of *EWSR1::FEV* (1/3) and in the single case harboring an *EWSR1::ETV4* fusion.

##### TP53 mutations are linked to poor outcomes in EwS

*TP53* was mutated in 7.1% (38/538) of EwS. Here, among those cases for which information on the fusion gene was available, *TP53* mutations were found in 8.6% (28/326) of *EWSR1::FLI1*, 9.7% (4/41) of *EWSR1::ERG*, and one of the three cases harboring *EWSR1::FEV* fusions (**Fig. 3**). Patients with *TP53*-mutated EwS were more likely to be found among those with metastatic disease (11.3.%; 17/150) than with localized disease (5.5%; 16/291; *P*<0.05). These patients were more prone to rapidly experiencing a new event of progression of disease, at mean of 18.4 months (±6.6) compared to 103.6 months (±10.5.1) for patients without *TP53* mutations (HR 3.63; 95% CI:1.5–8.77; *P*<0.01; **Supp. Fig. 3A**). Consequently, patients with *TP53* mutations had a shorter OS (20.7±6.1 *versus.* 109.3±10.25 months; HR 4.66; 95% CI:2.04–10.62; *P*<0.001; **Supp. Fig. 3B**, **Fig. 4**). In general, patients with *TP53*-mutated EwS were more likely to die due to disease [88.8% (24/27) *versus* 45.5% (131/288) of non-mutated cases; *P*<0.001], confirmed by logistic regression (OR=9.19, 95% CI: 2.287–27.963; *P*<0.001; **Table 3a**), which is in keeping with prior reports that identified *TP53* mutations as a negative prognostic marker^5,11,33–35^. In contrast to *STAG2* mutated cases, *TP53* mutations were more associated with EwS patients that were older than those with *TP53* wildtype tumors (mean age at diagnosis: 23.4±11.4 *versus* 19.3±12.7 years old, respectively; *P*=0.034; OR 1.02; 95% CI: 0.998–1.045; logistic regression *P*=0.072). **Table 3a**). When we segregated localized from metastatic categories, we observed that in both cases the presence of *TP53* mutations were associated with poor outcomes (**Tables 3b, c**). Given the small number of *TP53*-mutant cases, estimates should be interpreted with caution.

**Figure 3.**
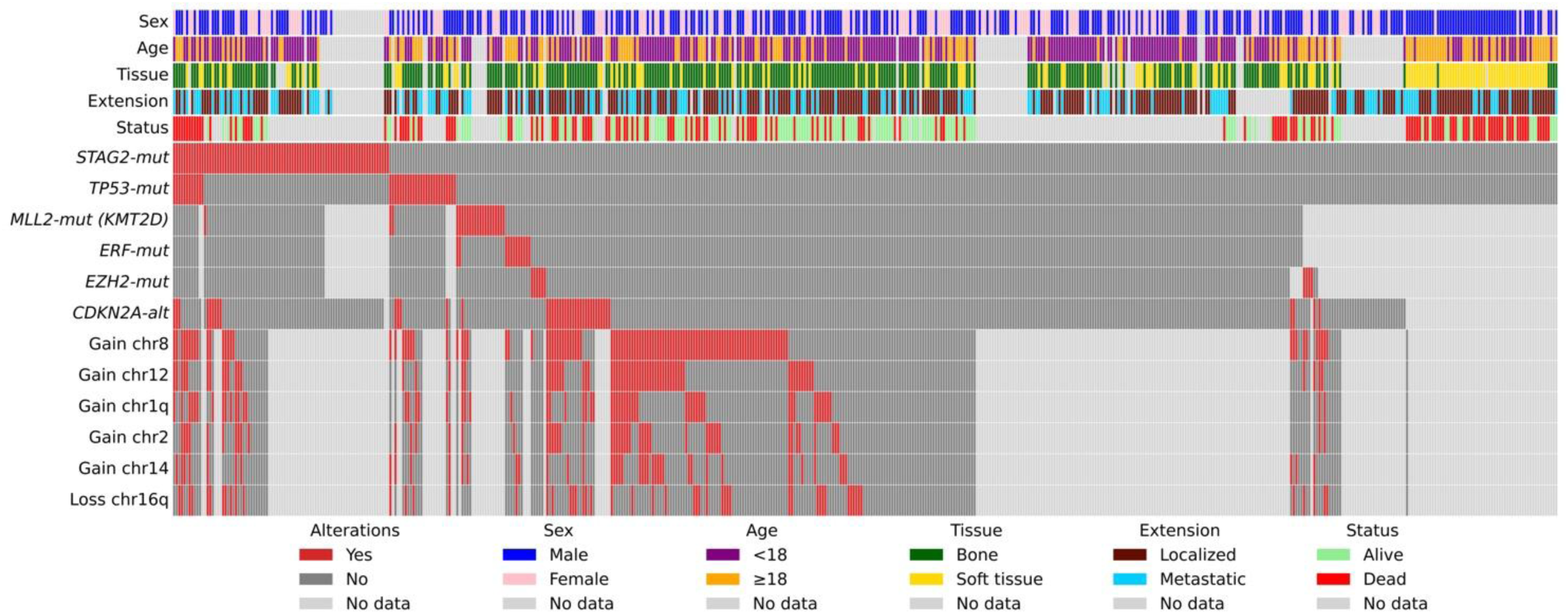
Oncoprint of the main genomic alterations in the study cohort. Columns represent individual tumor samples (*n*=538), and rows represent the clinicopathological variables and the most frequent alterations, as defined in the legend.

**Table 3.**
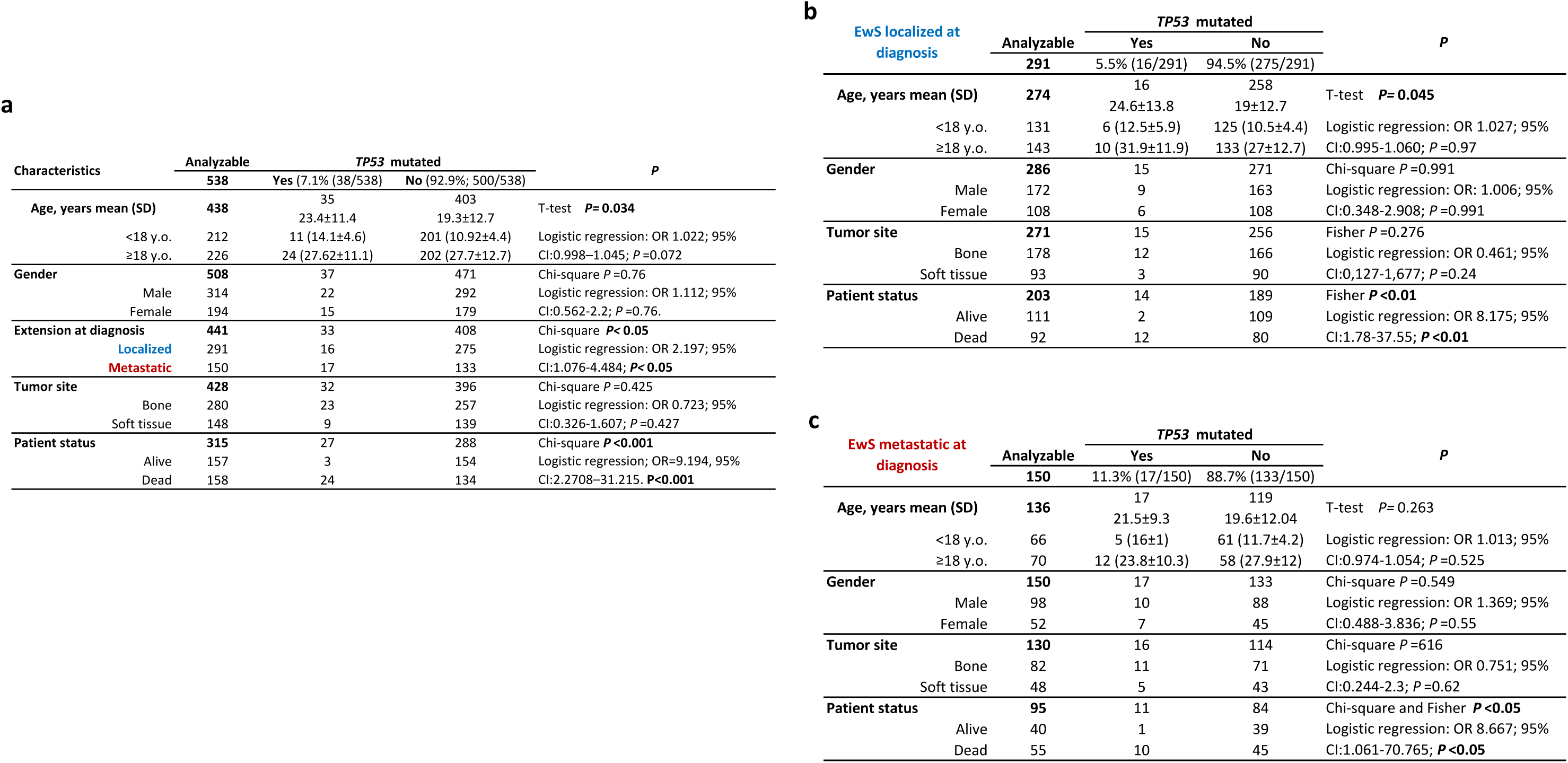
Clinicopathological implications of the presence of *TP53* mutations: **a)** Global study cohort; **b)** Localized cases. **c)** Metastatic cases.

##### Concomitant STAG2 and TP53 mutations are associated with fatal outcome

In our study cohort, 31.6% (12/38) of *TP53*-mutated EwS simultaneously harbored *STAG2* mutations, which is more than statistically expected by chance (*P*<0.05). Concurrent *TP53* and *STAG2* mutations were slightly more frequent in cases with metastasis at diagnosis (54.5% of cases; 6/11; *n*=441, *P*=0.14). Although the number of dual-mutant cases is small (*n*=12), the Fisher test analyzing deceased patients and concurrent *STAG2* and *TP53* mutations confirmed the association with fatal outcome (100%; 12/12; *P*<0.001), which is striking and consistent with prior observations^5^. EFS and OS data were only available for four dually mutated cases. On average, these four cases had shorter EFS (11±2.6 months) and OS (13.5±1.7 months) as compared to non-mutated cases (*n*=106; mean EFS of 102.3±10.3 months; *P*<0.05 and mean OS of 111± 9.4 months; *P*<0.05). In agreement with Tirode et al. (ref.^5^), a Cox model confirmed that patients harboring concomitant *STAG2* and *TP53* mutations had a higher risk for early progression (HR 6.63; 95% CI: 2.25–19.47; *P*<0.001) and a reduced survival probability (HR 7.05; 95% CI: 2.39–20.75; *P*<0.001) (**Suppl. Fig. 4**, **Fig. 4**). Yet, given the limited number of *STAG2*/*TP53*-double-mutant cases, these results need to be interpreted with caution.

**Figure 4.**
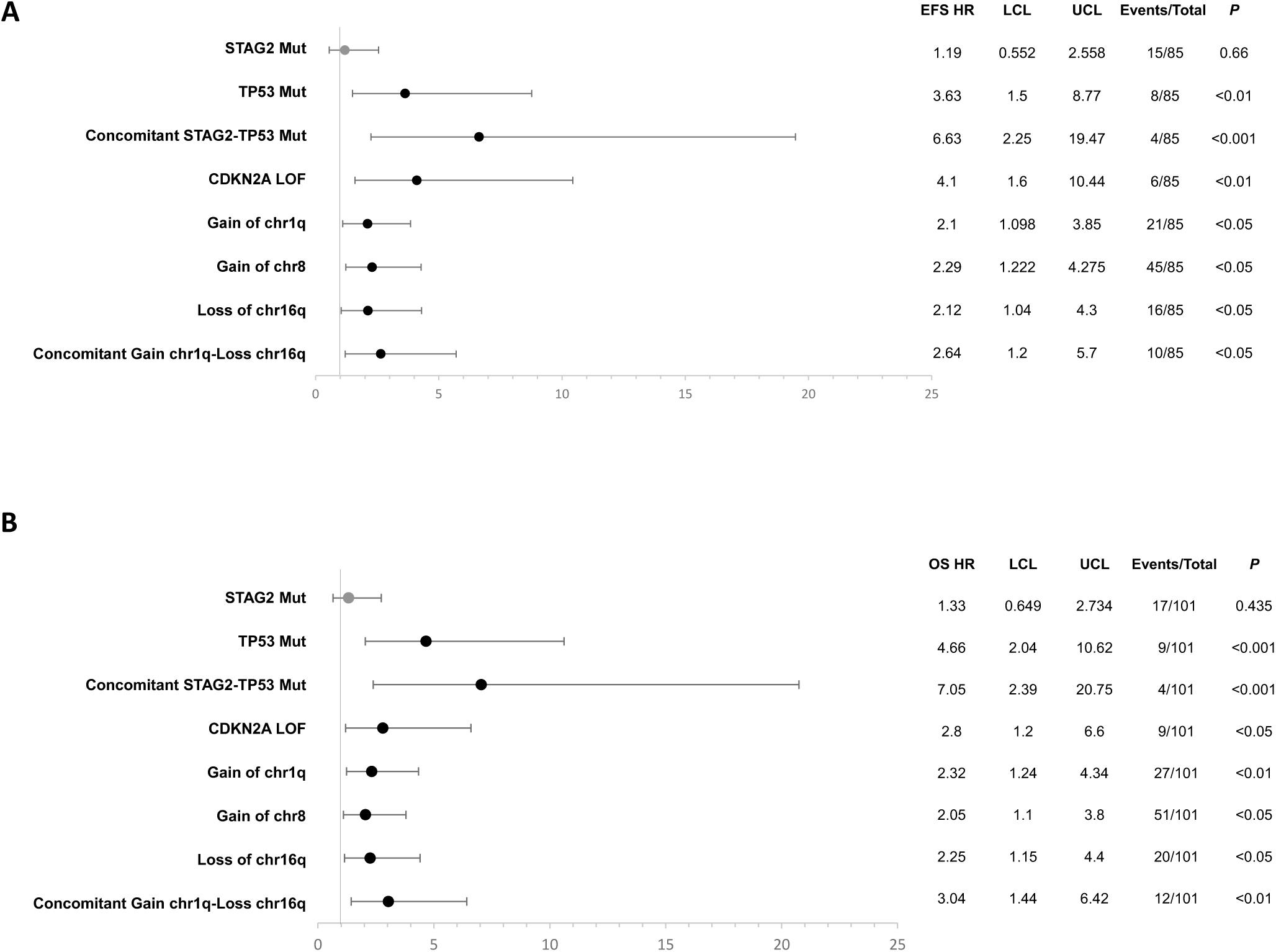
Risk of **(A)** reduced Event free survival (EFS) and **(B)** Overall survival (OS) of the main alterations described in EwS. HR: Hazard ratio. LCL: Lower confidence limit. UCL: Upper confidence limit.

Despite the small numbers, double-mutant cases exhibited no particular tendency for a major EWSR1::ETS fusion. Indeed, out of the 12 dually mutated cases, 10 harbored *EWSR1::FLI1* and two harbored an *EWSR1::ERG* fusion. *STAG2* was concomitantly mutated only with one *MLL2* mutated case (1/21) and with none of the *ERF* (consistent with previous observations^11,31^) or *EZH2* mutated cases. The same was observed in the case of *TP53* with two *MLL2* mutated cases (2/22) but being mutually exclusive with *ERF* or *EZH2* mutations in our cohort.

##### Clinicopathological features of less frequently mutated genes

The reduced number of *MLL2 (KMT2D), ERF* or *EZH2* mutated cases precluded reaching relevant values in most clinicopathological comparisons. However, we observed that *MLL2* mutations associated with survival (all 7 *MLL2*-mutated cases were annotated as alive; *P*<0.05). In the case of *ERF*, the presence of mutations was associated with older patients [26.7±12.4 years old (*n*=10) *versus* 17.8±12.1 years old (*n*=346); *P*<0.05; **Table 4**], as well as *EZH2* mutated cases [35.1±25.4 years old (*n*=9) *versus* 17.8±12 years old (*n*=348). *P*<0.001]. Tumors mutated in *EZH2* arose proportionally more frequently in soft tissues [5.8% (5/86) *versus* 1.5% (4/263); *P*<0.05; **Table 4**]. Only one *ERF* and three *EZH2* mutated cases had available EFS and OS information, precluding thorough review. As it was noted for *TP53*-mutant cases or *STAG2-TP53* dually mutant cases, given the small number of cases with mutations in these genes, estimates should be interpreted with caution.

**Table 4:** Clinicopathological implications of the most relevant molecular alterations. *P*-values refer to individual or univariate analyses. * Due to the reduced number of cases, statistics should be interpreted with caution.

| Molecular alteration | *n* | Clinicopathological implication | *P* |
| --- | --- | --- | --- |
| **STAG2 mutations** | 84 | More common in younger patients (15.8 vs. 20.2 y.o.)<br>Trend to associate with metastatic disease (17.3 vs. 11.3%) | <0.05<br>0.08 |
| **TP53 mutations** | 38 | More common in older patients (23.4 vs. 19.3 y.o.)<br>Associated with metastatic disease at diagnosis (11.3 vs. 5.5%)<br>Reduced EFS (18.4 vs. 103.6 months)<br>Reduced OS (20.7 vs. 109.3 months)<br>More commonly associated with lethal outcomes (88.8% vs. 45.5%) | <0.05<br><0.05<br><0.01<br><0.001<br><0.001 |
| **Concomitant STAG2+TP53 mutations** | 12 * | Reduced EFS (11 vs. 102.3 months)<br>Reduced OS (13.5 vs. 111 months)<br>Associated with lethal outcomes (100%) | <0.001<br><0.001<br><0.001 |
| **ERF mutations** | 12 * | More common in older patients (26.7 vs. 17.8 y.o.) | <0.05 |
| **EZH2 mutations** | 10 * | More common in older patients (35.1 vs. 17.8 y.o.)<br>Soft tissue prevalence (5.8 vs. 1.5%) | <0.001<br><0.05 |
| **CDKN2A alterations** | 43 | More common in older patients (23.7 vs. 18.2 y.o.)<br>Reduced EFS (23.7 vs. 54 months)<br>Reduced OS (24 vs. 60.4 months)<br>More commonly associated with lethal outcomes (72 vs. 39.9%) | <0.05<br><0.05<br><0.05<br><0.01 |
| **Concomitant CDKN2A+TP53 alterations** | 7 * | Associated with lethal outcomes (100%) | <0.01 |
| **Gain of chr1q** | 61 | Trend to associate with male (29.4 vs. 18.8%)<br>Associated with metastatic disease at diagnosis (35 vs. 19.6%)<br>Reduced EFS (33.3 vs. 48.2 months)<br>Reduced OS (41 vs. 68 months)<br>More commonly associated with lethal outcomes (37.1 vs. 16.9%) | 0.07<br><0.05<br><0.05<br><0.05<br><0.001 |
| **Gain of chr8** | 124 | Reduced EFS (42 vs. 63 months)<br>Reduced OS (58.2 vs. 64.1 months)<br>More commonly associated with lethal outcomes (60 vs. 40%) | <0.05<br><0.05<br><0.05 |
| **Gain of chr20** | 57 | Reduced EFS (34.7 vs. 57.1 months) | <0.05 |
| **Gain of chr14** | 46 | Reduced EFS (30 vs. 57 months) | <0.05 |
| **Loss of chr16q** | 53 | Bone tumor prevalence (88.5%)<br>Reduced EFS (30.7 vs. 56.7 months)<br>Reduced OS (38.8 vs. 66.4 months)<br>More commonly associated with lethal outcomes (60.8 vs. 38.5%) | <0.05<br><0.05<br><0.05<br><0.01 |
| **Concomitant gain of chr1q + loss of chr16q** | 25 | Reduced EFS (22.2 vs. 55.8 months)<br>Reduced OS (30.9 vs. 65 months)<br>More commonly associated with lethal outcomes (69.6 vs. 40.6%) | <0.05<br><0.05<br><0.01 |
| **Concomitant STAG2 mut + gain of chr1q** | 13 | Bone tumor prevalence (85%)<br>Associated with metastatic disease at diagnosis (69%)<br>More commonly associated with lethal outcomes (83.3%) | <0.05<br><0.05<br><0.05 |
| **Concomitant STAG2 mut + gainChr1q + lossChr16q** | 8 * | Bone tumor prevalence (85.7%)<br>Associated with metastatic disease at diagnosis (57.1%)<br>More commonly associated with lethal outcomes (85.7%) | <0.05<br><0.05<br><0.01 |
| **Concomitant TP53 mut + gain chr8/1q/20/loss16q** | 7-16 * | Bone tumor prevalence (81.2, 90, 66.7 and 100%, respect)<br>TP53 mut+gain chr8,-1q,-20 - Metastatic disease (19.5, 60 and 66%)<br>TP53 mut+loss chr16q - Localized disease (85.7%)<br>More commonly associated with lethal outcomes (88, 91, 91 and 87.5%, respect) | <0.05<br><0.05<br><0.05<br><0.05 |
| **Concomitant CDKN2Aalt + gain chr8/chr12/loss16q** | 24, 17 and 16 | Male predominance (63.6, 70.6%, NS)<br>Associated with localized disease at diagnosis (75, 82%, NS)<br>Bone tumor prevalence (83.3, 94 and 85%)<br>More commonly associated with lethal outcomes (64, 69 and 83%) | <0.01<br><0.01<br><0.01<br><0.01 |

### Loss of *CDKN2A*

Recurrent *CDKN2A* alterations are well-known in EwS, usually detected as focal deletions, but also as point mutations^1,36^. We considered both deletions and point mutations as loss-of-function (LOF) alterations of *CDKN2A.* Loss of *CDKN2A* was present in 9.1% (43/473) of EwS cases, within the range described in previous reports on 16 clinical studies (6–32%)^11,36^. In our study cohort, loss of *CDKN2A* was associated with a higher age at diagnosis (22.7±1.1 *versus* 18.2±12.6 years; *n*=373; *P*<0.05), reduced EFS time [23.7±36 (n=6) *versus* 54±45.7 (n=79) months; *P*<0.05. HR 4.1; 95% CI:1.6–10.44; *P*<0.01], reduced OS [24±28.3 (*n*=9) *versus* 60.4±48.6 (n=101) months; *P*<0.05. HR 2.8; 95% CI:1.2–6.6; *P*<0.05] and with higher mortality rates [72% (23/32) EwS patients with *CDKN2A* loss died *versus* 39.9% (87/218); *n*=250; *P*<0.01; **Fig. 4**]. This was confirmed by binary logistic regression (OR 3.85 95% CI: 1.7–8.8; *P*<0.001).

*CDKN2A* alterations co-occurred with *STAG2* mutations in 9/43 cases, with *TP53* mutations in 7/43 cases and with *MLL2* in 1/40 cases, but with neither *ERF* nor *EZH2* (**Fig. 3**). Conversely, 20% of *TP53*-mutated tumors (7/35) also carried *CDKN2A* alterations (*P*<0.05). In contrast, co-occurrence with *CDKN2A* alterations was observed in only 11% of *STAG2*-mutated tumors (9/81) (*P*=0.49), *MLL2* (*P*=0.4). All *CDKN2A* alterations co-occurring with *STAG2* mutations (derived from refs^3,4,7^) were focal deletions.

Co-occurrence of *CDKN2A* with *STAG2* or *MLL2* alterations was not strongly associated with any clinical feature. However, concomitant alterations in *TP53* and *CDKN2A* tended to be (*n*=365; *P*=0.064) more frequently detected in soft tissue EwS (57%; 4/7; *n*=92) than in bone EwS (13%; 3/23; *n*=273). Importantly, 6/6 of patients (100%) with concomitant *STAG2* and *CDNK2A* mutations and for which vitality status data were available died of the disease (*P*<0.01), supporting previous observations^11^. Cox regression analyses were not possible since EFS or OS were not available for the 7 concomitantly mutated cases.

Of note, neither *ERF* nor *EZH2* mutations were found in cases with *STAG2*, *TP53* or *CDKN2A* alterations. In the case of *ERF* mutations, this coincided with a recent report describing their potential mutual exclusivity with *TP53*, *CDKN2A* and *STAG2* mutations^11^ (**Fig. 3**).

#### Copy number variations (CNVs)

CNVs are alterations characterized by the loss or gain of DNA sequences that are larger than 50 bp^37^. In our cohort, CNV data were available for 242 cases. Chr8 was gained in 51.2% (124/242), chr12 in 29.3% (71/242), chr1q in 25.2% (61/242) and chr20 in 23.6% (57/242) of cases. Additionally, loss of chr16q was found in 21.9% (53/242). These proportions are in line with the literature [gains of chr8: 26.9 (ref.^8^) – 83.3% (ref.^7^); chr12: 16.6 (ref.^7^) – 33%(ref.^38^); chr1q:10.2 (ref.^11^) – 31% (ref.^2,7^); chr20: 10–20% (ref.^1^), loss of chr16q: 12.2% (ref.^11^) – 21% (ref.^39^)]. We also found a high frequency of gains in chr2 in 21.9% (53/242) and in chr14 in 19% (46/242) of cases, previously reported in ∼9% (ref.^11^) – 25% (ref.^1^) and 9% (ref.^39^) – 11% (ref.^11^) of EwS cases, respectively, although without clinical relevance, except for gains in chr14q, which correlated with poor progression free survival (ref.^11^), a trend that we also observed in our study cohort (30.4±25.4 *versus* 57.6±48.2 months. *P*<0.05; HR 1.88; 95% CI: 0.96−3.7; *P*=0.065). We found that many of these CNVs were frequently concomitant (**Supp. Table 2**), particularly the co-occurrence of gains of chr8 and chr12 in 52 cases (*P*<0.001). Among the 242 cases analyzed, 178 (73.5%) showed CNV on any of the mentioned chromosomes (**Fig. 3**). To explore whether these CNV were predictive factors for clinical outcomes, we analyzed each of them both individually and in combination in the context of clinical parameters.

First, we observed that none of the CNVs were associated with a specific type of oncogenic fusion. While gains of chr8 were similarly distributed in terms of age at diagnosis, sex, tumor localization (bone/soft tissue), or disease stage at diagnosis, they were more frequent among those patients experiencing reduced EFS (42±43.5 *versus* 63±45.9 months; HR 2.29; 95% CI:1.22−4.27; *P*<0.05), reduced OS (58.2±53.5 versus 64.1±42.5; HR 2.05; 95% CI:1.1−3.8; *P*<0.05. **Fig. 4**) and consequently, those who died due to the disease [60% (60/100) *versus* 40% (40/100); *P*<0.05. **Table 4**). Neither the gains of chr2 nor chr12 were associated with any of the analyzed factors. However, gains in chr20 associated with reduced EFS (34.7±32.4 *versus* 57.1±47.9; *P*<0.05. HR 1.96; 95% CI:1.04–3.7; *P*<0.05). Gains in chr1q tended to associate with male patients [29.45% (43/146) *versus* 18.8% (17/90); *P*=0.07] and were associated with the presence of metastasis at diagnosis (35%; 28/80) as compared to localized disease (19.6%; 30/153; *P*<0.05), reduced EFS (33.3±30.3 *versus* 58±48.2 months; *P*<0.05. HR 2.1; 95% CI:1.1−3.9; *P*<0.05), reduced OS (41±35.4 *versus* 68±50.5 months; *P*<0.05. HR 2.32; 95% CI:1.24−4.34; *P*<0.01; **Fig. 4**), and consistently linked to those patients who died of the disease [37.11% (36/97) *versus* 16.9% (22/130); *P*<0.001], in agreement to what was previously described^1,2,11,40^. Loss of chr16q was associated with bone tumors [88.5% (46/52) cases with loss of chr16q arose in bone; *P*<0.05)], reduced EFS (30.7±37.9 *versus* 56.7±46 months; *P*<0.05; HR 2.12; 95% CI:1.04–4.3; *P*<0.05), reduced OS (38.8±34.2.4 *versus* 66.4±50 months; *P*<0.05; HR 2.25; 95% CI:1.15–4.4; *P*<0.05; **Fig. 4**), and death of disease [60.8% (31/51) patients with loss of chr16q died of disease *versus* 38.5% (69/179); *P*<0.01]. We found a correlation of cases with gain of chr1q and cases with loss of chr16q (*P*<0.01), as previously reported^40^. Those cases with concomitant alterations (25/242), experienced reduced EFS time (22.2±19 *versus* 55.8±46.7 months; *P*<0.05; HR 2.64; 95% CI:1.2–5.7; *P*<0.05), reduced OS (30.9±21.7 *versus* 65±49.5 months; *P*<0.05; HR 3.04; 95% CI:1.44–6.42; *P*<0.01; **Fig. 4**) and were more prone to die [69.6% (16/23) *versus* 40.6% (84/207); *P*<0.01; **Table 4**, **Fig. 3**). However, we did not find clinical association for the combined gain of chr8 and chr12, present in 52 patients.

We performed a multivariate logistic regression to evaluate the CNV most powerful predictor of mortality and observed that gain of chr1q was the most significant predictor of poor outcome in agreement with previous studies^40,41^ (OR 2.24; 95% CI: 1.2–4.2; *P*=0.013) followed by loss of 16q (OR 1.98; 95% CI: 1.01–3.8; *P*=0.04) and gain of chr8 (OR 1.73; 95% CI: 0.99–2.98; *P*=0.05).

#### Impact of CNV and SNV combinations on clinical features

Once we performed a comprehensive analysis of the clinical implications of the main somatic genomic alterations individually, we found that the most common CNVs and some of the most frequent recurrent mutations in EwS could coexist and be associated with clinical features.

When combining frequently altered genes with CNV, we detected that 85% (11/13) of cases harboring *STAG2* mutations and gains of chr1q arose in bone (*n*=237; *P*<0.05), being 69% (10/13) of these cases metastatic at diagnosis (*n*=233; *P*<0.05), and 83.3% (10/12) died of the disease (*n*=230; *P*<0.05). When we added concomitant loss of 16q to these comparisons, some differences became more significant. *STAG2* mutations together with gains of chr1q and loss of chr16q were associated with bone tumors in 85.7% of cases (6/7; *n*=237; *P*<0.05), with the presence of metastasis at diagnosis in 57.1% (4/7; *n*=233; *P*<0.05) and with lethality in 85.7% of cases (6/7; *n*=230; *P*<0.01; **Table 4**).

Likewise, *TP53* mutations in combination with gains of chr8, gains of chr1q, or loss of chr16q were more frequently detected in EwS of bone [81.2% (13/16), 90% (9/10) and 100% (7/7) of cases, respectively) as compared to cases arising in soft tissue (*n*=237; *P*<0.05 for all comparisons). These conjunctions are associated with death of disease in 88.2% (15/17), 91% (10/11) and 87.5% (7/8) of cases, respectively (*n*=230; *P*<0.05). Combination of *TP53* mutations and gains of chr20 was also associated with lethality in 91% (10/11) of cases (*n*=230; *P*<0.05). Among EwS patients with concomitant *TP53* mutations and CNV, we observed coincidences with gain of chr8 in 19.5% (8/41) of metastatic cases, whereas they were found in only 10.3% (8/78) of localized cases (*n*=233; *P*<0.05). In the case of concomitant *TP53* mutations and gains of chr1q, 60% (6/10) were already diagnosed with metastatic disease (*n*=233; *P*<0.05). In contrast, *TP53* mutated cases with loss of chr16q were more frequently observed in localized cases (6/7, *P*<0.05), although the limited number of cases requires cautious interpretation.

Significant associations linked *CDKN2A* loss with gains of chr8 in 75% (24/32) of cases (*P*<0.01); with gains of chr12 in 53% (17/32) of cases (*P*<0.01); and with loss of chr16q in 41% (13/32) of cases (*P*<0.05; **Fig. 3**). We found that most of EwS with *CDKN2A* alterations in combination with gains of chr8 arose in males (63.6%; 14/22; *n*=235; *P*<0.01) as localized disease (75%; 18/24) in bone (83.3%; 20/24) (*n*=236; *P*<0.01), of which 64% (14/22) died due to the disease (*n*=229; *P*<0.01).

Also, EwS with *CDKN2A* alterations and gains in chr12 were more frequent in males (70.6%; 12/17; *n*=235; *P*<0.01), arose in bone in 94% (16/17) of cases (*n*=236; *P*<0.01), 82% (14/17) of which were localized at diagnosis (*n*=232; *P*<0.01), and 69% (11/16) of these patients died (*n*=229; *P*<0.01).

We also observed relative predominance of male patients (83.3%; 10/12; *n*=235; *P*=0.052) when *CDKN2A* alterations coincided with gains of chr1q, as well as bone localization (75%; 9/12; *n*=236; *P*=0.092). Although low numbers, it should be noted that co-occurrence of *CDKN2A* alterations and gain of chr20 was more frequently observed in males (5/6, *P*=0.7) and in tumors arising as localized disease in bone (5/7, *P*=0.7). An association with bone origin occurred when *CDKN2A* alterations coincided with loss of chr16q in 85% (11/13) of cases (*n*=236; *P*<0.01). Also, *CDKN2A* alterations in combination with chr16q loss were associated with death of the disease (83%, 10/12 of patients; *n*=229; *P*<0.05). 100% of patients (4/4) with concomitant *CDKN2A* alterations, gain of chr1q and loss of 16q were male, with bone tumors (4/4); metastasis at diagnosis in 75% (3/4) and 100% of mortality (3/3).

From the very few cases with CNV data and information on the other most recurrently mutated genes, we observed the following: 3/8 *EZH2*-mutated cases, 2/7 *ERF*, and 2/3 *MLL2* mutated cases also show gain of chr8; 1/8 *EZH2* mutated cases, 1/3 *MLL2*, also show gain of chr12; 1/7 *ERF* mutated cases and 2/3 *MLL2*, also show gain of chr1q; 2/8 *EZH2* mutated cases, 2/7 *ERF* and 2/3 *MLL2*, also show gain of chr20; 1/8 *EZH2* mutated cases and 1/7 *ERF,* also show loss of 16q **(Fig. 3)**. The reduced number of cases precludes performing statistical analysis for clinical effects, although certain level of genomic instability may be inferred from these concomitances considering the global low frequency of genomic alterations in EwS.

Finally, we performed a global multivariable logistic regression to identify factors associated with EwS-specific mortality. *TP53* mutation status (OR 7.09; 95% CI: 1.92–26.1; *P*=0.003) and gain of chr1q (OR 1.98; 95% CI:1–3.89; *P*=0.047), emerged as independent predictors of mortality, indicating a markedly increased risk of death from disease among affected cases. Borderline associations were observed for *CDKN2A* alterations (OR 2.44; 95% CI:0.99–6.03; *P*=0.053). By multivariate Cox model we observed that mutated *TP53* (HR 2.75; 95% CI:1.03–7.34; *P*=0.044) and *CDKN2A* alterations (HR 3.53; 95% CI:1.14–10.9; *P*=0.028) are the main predictors of a reduced event free survival and in the case of *TP53* mutations, also a reduced overall survival (HR 3.29; 95% CI:1.19–9.1; *P*=0.022).

## METHODS

### Source of EwS samples and associated clinicopathological information

#### Aggregated samples from published studies (‘Study cohort’)

We included sequencing data from 550 EwS samples [*n*=132, ref.^5^; *n*=99, ref.^4^; *n*=92, ref.^6^; *n*=83 unique (not included in the study from Nacev et al.), ref.^3^; *n*=72, ref.^7^; *n*=52, ref.^8^; *n*=20; ref.^9^] from different published studies^3–9^. We excluded 4 cases being originally classified as EwS, but showing *FUS::NFATc2* (ref.^7^), *ETV6::NTRK3* (ref.^7^), *CIC::FOXO4* (ref.^7^) and *EWSR1::NFATc2* (ref.^4^) fusions according to the details provided, thus representing other tumor entities^42,43^. Other 4 cases were discarded for no presenting any of the known fusions^7^ and other 4 cases for not being evaluable for mutations (*n*=538). Data censoring was done on March 1^st^ 2024.

### Data processing

#### Clinicopathological data

Clinicopathological data were extracted from the original reports and associated metadata, being available for most patients included in the study. We analyzed the age at diagnosis (*n*=438), sex (*n*=508), tissue of origin (*n*=428), primary or secondary origin of the tumor sample (*n*=411), extension at diagnosis (*n*=441) and status of the patient (*n*=315). EFS and OS data were consistent and available for 85 and 103 patients, respectively.

#### Mutational data

Depending on the study, mutational data were derived from WGS^5–7^; WES^6,8^; SNP arrays^6^ and/or targeted sequencing^7^, including the MSK-IMPACT (Memorial Sloan Kettering-Integrated Mutation Profiling of Actionable Cancer Targets) workflow^3,4,44^. Some studies used the same MutSig algorithm to attribute significance to DNA mutations by identifying genes that were mutated more often than expected^4,6,8^. We adopted the conversion and normalization of the respective raw data and directly used the reported mutational information for the given samples from those studies.

#### Fusion types included in the study

We included information about the FET::ETS fusions for 374 patients: 326 EWSR1::FLI1 (*n*=93, ref.^5^; *n*=77, ref.^3,4^; *n*=69, ref.^3^; *n*=51, ref.^7^; *n*=20, ref.^6^; *n*=15, ref.^8^; *n*=1, ref.^4^), 41 EWSR1::ERG (*n*=15, ref.^3,4^; *n*=10, ref.^3^; *n*=9, ref.^5^; *n*=4, ref.^6^; *n*=3, ref.^7^), 3 EWSR1::FEV (*n*=2, ref.^7^; *n*=1, ref.^4^); 2 FUS::FEV (ref.^4^); 1 EWSR1::ETV4 (ref.^4^) and 1 EWSR1::ETV1 (ref.^5^). Most of gene fusions were primarily estimated by FISH as part of the routine protocol, and depending on the study, *FET::ETS* rearrangements were detected also by WGS^5–8^, and fusion transcripts were confirmed by qRT-PCR^5^, RNAseq^6,7^, or as a part of a pipeline such as that from the MSK-IMPACT^3,4,44^.

#### Copy Number Variations (CNVs)

Likewise, we used the information on copy number variations provided in the corresponding articles^3,5–7^ assuming conversion and normalization of the respective raw data. CNV information was available for 242 EwS samples (*n*=105, ref.^5^; *n*=91, ref.^4^; *n*=21, ref.^3^; *n*=18, ref.^6^; *n*=7, ref.^7^).

### Statistical analysis

Statistical analyses were performed using IBM SPSS Statistics v29.0.2.0. All analyses were conducted on the available cases for each variable, without imputation of missing data. Because the aggregated cohort was derived from heterogeneous published studies and sequencing platforms, genomic alterations were harmonized into binary variables, including mutated *versus* non-mutated status for recurrently altered genes and presence *versus* absence of copy-number gains or losses for CNVs.

Descriptive statistics were used to summarize clinicopathological and molecular variables. Categorical variables were expressed as absolute numbers and percentages, whereas continuous variables were reported as mean ± standard deviation, unless otherwise specified. Associations between categorical variables were evaluated using Pearson’s chi-square test or Fisher’s exact test when the analyzed subgroup included fewer than 20 cases or when at least one expected cell count was below 5. Comparisons of continuous variables between two groups were performed using Student’s t-test or the Mann–Whitney U test, depending on data distribution and group size. For comparisons involving more than two groups, one-way ANOVA was used when appropriate.

Binary logistic regression models were used to evaluate associations between selected clinicopathological or molecular variables and binary outcomes, including mutation status, tissue of origin, presence of metastatic disease at diagnosis, and EwS-specific mortality. Results were expressed as odds ratios (ORs) with 95% confidence intervals (95% CI). Model calibration was assessed using the Hosmer–Lemeshow goodness-of-fit test, considering values >0.05 as indicative of adequate fit. Multivariable logistic regression models were used to identify independent predictors of EwS-specific mortality, incorporating clinically relevant variables and genomic alterations that showed significant or borderline associations in univariable analyses.

EFS and OS analyses were restricted to cases with consistent and evaluable follow-up information. EFS was defined as the time from diagnosis to disease progression, relapse, or death from disease, whereas OS was defined as the time from diagnosis to death from EwS or last follow-up. Cases from datasets with incomplete, inconsistent, or non-comparable survival annotation were excluded from survival analyses. Cox proportional hazards regression was used to assess the impact of clinicopathological and genomic variables on EFS and OS. Hazard ratios (HRs) were reported with 95% CI.

Given the limited number of cases harboring some molecular alterations, including rare FET::ETS fusion types, *TP53*-mutant cases, *STAG2/TP53* double-mutant cases, and less frequently mutated genes such as *MLL2*, *ERF* and *EZH2*, statistical estimates from these subgroup analyses were interpreted with caution. The analyses conducted as part of the study were exploratory in nature. Therefore, the p-values derived from all statistical tests -which were exclusively two-tailed tests-were interpreted merely as an indicator of significance. Oncoprint visualizations were generated using Python v3.9.

## DISCUSSION

This study integrates molecular and clinicopathological data from the largest pooled EwS cohort reported to date, confirming observations from smaller series and providing novel insights previously unattainable with narrower datasets. Analytic integration enabled the detection of associations that were underpowered in the original individual studies.

We assembled a cohort of 538 EwS patients by compiling molecular and clinical data from seven published sequencing studies^3–9^. The distribution of clinicopathological features was consistent with previous reports^3–7^. However, both the mean age at diagnosis (19 years) and the proportion of soft-tissue tumors (34.6%) exceeded expectations (∼15 years and 10–15% of soft-tissue origin^1^, respectively). In part, this reflects the association between age and tissue of origin: older patients more frequently developed soft-tissue tumors, consistent with prior studies^45–47^. We observed that the largest series^3,4^ incorporated in our study cohort -representing 33% (177/538) of patients-was derived from non-specific pediatric centers, including mostly adult patients with a mean age of almost 28 years old and a consistent increased percentage of soft tissue EwS (49.1%; 86/175). Both facts affected the global representation in the study cohort. Also, technical challenges in obtaining adequate DNA from bone biopsies may have biased the series toward soft-tissue tumors, artificially raising the mean age. As this cannot be confirmed for each study, we acknowledge that this is a potential limitation of our pooled analysis.

In our aggregated cohort, younger patients (here mean 19 years) were more likely to present with metastases, consistent with prior findings grouping the higher proportion of patients with metastatic disease at diagnosis in the range of 15–19 years of age^47^. However, our analyses showed that this was not directly linked to mortality, as the mean age of deceased patients was relatively higher (23.6 years). This interpretation should be considered with caution due to the enrichment in older patients derived from Nacev and Dermawan series, which largely contributes with vital status data and may bias the associations with patient outcome. According to our data, prognosis appeared to be associated with combinations of factors such as tissue of origin, sex, and age since soft-tissue tumors were more common in older males and associated with worse outcomes, or individual but very relevant factors such as the presence of metastasis at diagnosis.

The prognostic significance of FET::ETS fusion type remains controversial^21,22^. In our 374 evaluable cases, fusion types were represented at expected frequencies and showed no association with clinical features, consistent with part of the literature^1,10,11,40^.

As previously reported^3–10,31^, *STAG2* and *TP53* were the most frequently mutated genes (15.6% and 7.1%, respectively), but we also catalogued other less frequent mutated genes such as *MLL2 (KMT2D), ERF* or *EZH2*. We excluded *TTN* due to its length and high background mutation rate in most cancer studies^24^. Also two very large genes, with repetitive sequences, that were frequently mutated across multiple sequencing studies with no functional driver roles, *MUC6* and *AHNAK2,* were discarded due to the lack of validation in the original work^6^. *KDR* was excluded because nearly all cases shared a likely benign germline variant (Q472H, c.1416A>T) restricted to one study^9^.

Younger patients had a higher frequency of *STAG2* mutations, which showed a statistical trend for an association with metastatic presentation, in agreement with previous reports^5,6,10,13,14^. However, the presence of *STAG2* mutations did not predict mortality. In contrast, *TP53* mutations were strongly associated with reduced EFS, reduced OS and lethality, supporting its role as a negative prognostic marker in EwS^5,33–35^. *TP53* mutations were associated with older patients, consistent with the age-related increase of *TP53* mutation frequency observed in other cancers^48–53^. Notably, 31.6% of *TP53*-mutant cases also harbored *STAG2* mutations; of these, 70% were metastatic, 80% originated in bone, and all patients died, confirming the poor prognosis of this combination^5^. This contrasts with Gillani *et al*.^10^, who reported a stronger prognostic impact of *STAG2* loss alone and no strong association with patient age. Since that study^10^ only included patients with localized disease at diagnosis, we separated localized *versus* metastatic cases and observed that the association between *STAG2* mutations and younger age was particularly strong in localized cases, confirming the initial discrepancy described for our whole cohort. However, our findings concur with those of Gilliani *et al.*^10^ when segregating the analysis of clinical implication of *STAG2* mutations in two groups of patients: <18 *versus* ≥18 years old, since 89% of patients included in their study were younger than 18 years old, we selected the same age range and obtained a significant association between the presence of *STAG2* mutations and fatal outcome in our cohort [66.7% (10/15) of patients harboring *STAG2* mutations died, in comparison with 36% (35/97) of non-mutated patients. *P*<0.05]. Yet, we did not find this association when restricting our analysis to only patients with localized disease and less than 18 years old.

In our study cohort, the multivariable logistic regression analysis indicated that *TP53* mutations were the most powerful predictor of poor outcome. Co-occurring alterations involved *STAG2* or *TP53* together with *MLL2 (KMT2D),* or some of the excluded genes, such as *MUC6* or *AHNAK2* mutations. Although no strong associations emerged, the co-occurrence of diverse low-frequency alterations with *STAG2* or *TP53* suggests an increased tumor mutational burden (TMB). On the one hand, an association between *STAG2* and *TP53* mutations has been previously reported in EwS and has been linked to elevated TMB^54^. On the other hand, the concomitant genes, despite being described as “frequently” mutated in EwS, each display a mutational frequency of <6% across cases. Therefore, the presence of more than one of these mutations in the same patient, in addition to *STAG2* or *TP53,* appears unlikely to be coincidental and may instead reflect a broader pattern of genomic instability.

Although raw sequencing data were not always available, we were able to explore this hypothesis in a subset of 20 cases, including two cases with *STAG2* mutations and four cases with *TP53* mutations. In both groups, TMB was higher; particularly outstanding for *TP53* mutations (*P*<0.05). These findings suggest that the relationship between TMB and these mutations warrants further investigation in future studies.

Loss of *CDKN2A*, a common EwS alteration (>5%)^1^, has been controversially associated with prognosis^5–7,34,55,56^. We considered *CDKN2A* alterations as both focal deletions and SNVs, and observed associations with older age, reduced EFS, OS time, and mortality. Co-occurrence of *CDKN2A* alterations and *TP53* mutations, particularly in soft-tissue tumors, was consistently associated with fatal outcome (100%). This supports the context-dependent role of *CDKN2A* alterations in EwS proposed by Paragji et al.^36^. Although we identified concomitant *STAG2* mutations and *CDKN2A* alterations in nine cases, all were characterized by focal *CDKN2A* deletions. Therefore, these findings do not contradict the mutual exclusivity between *STAG2* and *CDKN2A* mutations proposed by Tirode et al.^5^.

Analysis of CNVs in 242 patients revealed frequent gains of chr8 (51.2%), chr12 (29.3%), chr1q (25.2%) and chr20 (23.6%), and losses of chr16q (21.9%), consistent with ranges reported in prior studies^1,2,5,7,8,38,39^. Co-occurrence of chr1q gain and chr16q loss, often due to unbalanced t(1;16)^1^, was associated with metastasis and lethality^5,54^. Individually, gains of chr8 and chr1q, as well as loss of chr16q, also correlated with reduced time of EFS, OS and mortality^2,5,54^. Interestingly, it has been recently reported that aggregated genome-wide CNV alterations may represent a better predictor of EwS patient outcome than individual CNVs^40^. We observed that interactions between CNVs and certain mutated genes had prognostic implications. *STAG2* mutations combined with chr1q gain were linked to bone tumors and metastasis at diagnosis; the combination of *STAG2* mutation, chr1q gain and chr16q loss was lethal in 86% of cases. *TP53* mutations coinciding with chr8, chr1q or chr20 gains, or chr16q loss, predicted bone origin and mortality in >87% of cases, consistent with the poor prognosis observed for these combinations in complex-karyotype AML^51^. In that case, *TP53* mutations were correlated with older age, genomic complexity, and specific CNV, including gains in chr8 and losses in chr16q^51^, supporting our results and suggesting a level of genomic complexity that warrants further exploration.

*CDKN2A* loss together with chr8 gain and chr12 gain, were associated with 64% and 69% mortality, respectively, while co-occurrence with chr16q loss reached 83% mortality, highlighting chr16q loss as a strong negative marker^39,57^. Bone origin was observed in 83–94% of these cases, most of them being localized at diagnosis.

In line with the hypothesis that secondary alterations in EwS are non-random and biologically relevant^36,58^, we detected *ERF* and *EZH2* as genes frequently mutated in EwS and being mutually exclusive with respect to the most relevant ones: *STAG2*, *TP53* and *CDKN2A*, in agreement with previous observations^11,31^. On the one hand, *ERF* mutations may act as cooperating alterations that potentiate the transcriptional activity of EWSR1::FLI1. By impairing ERF repressor function, inhibition at ETS binding sites is reduced, allowing for enhanced EWSR1::FLI1-driven transcriptional programs^59^, defining a mechanistic role in EwS. On the other hand, *EZH2* is a target of EWSR1::FLI1 that contributes to tumor growth and metastasis^60^. *In vitro* studies assessing whether *EZH2* mutations confer gain-of-function effects -some of which seem to be currently in progress^32^-will be critical to clarify their role in EwS progression.

Although our pooled analysis represents the largest molecular and clinicopathological study in EwS to date, it has several limitations: First, sequencing platforms, depth and CNV-calling algorithms varied across cohorts, introducing technical heterogeneity. Second, raw sequencing data were not available for unified reprocessing, limiting harmonization. Third, treatment regimens and supportive care differed between studies and could not be controlled. Fourth, some genomic alterations were present in only a few cases, reducing statistical power. Fifth, in most cases, the vital status was not accompanied by average follow-up, so these data should be interpreted with caution. Finally, ethnic and geographical representation across the cohorts was uneven, particularly regarding variants enriched in specific populations.

In summary, by integrating genomic and clinicopathological data from the largest EwS cohort to date, we uncovered associations that hitherto remained obscured in underpowered smaller studies. Younger patients were more likely to present with metastases, but this was not directly predictive of mortality. *STAG2* mutations correlated with young age and relative predominance of metastatic presentation without being necessarily associated with lethality. In contrast, *TP53* mutations were more frequently found in older patients and strongly correlated with death, especially in combination with loss of *STAG2, CDKN2A*, or specific CNV. *CDKN2A* loss also contributed to mortality, particularly when combined with *TP53*. CNVs in chr8, chr1q, and chr16q further stratified prognosis, with chr1q gain and chr16q loss confirmed as especially adverse markers. Not all clinical patterns can be explained by these alterations, suggesting additional drivers, including epigenomic or treatment-related factors, warrant further study.

## Data Availability

All data produced in the present study are available upon reasonable request to the authors

## AUTHOR CONTRIBUTIONS

L.R.-P. and T.G.P.G. designed the study, curated the study cohort, carried out statistical analyses, drafted the paper and prepared the figures. C.H., A.R. J.D.-M., F.C.-A., U.D., and E.d.A. helped in data interpretation and provided statistical, clinical and pathological guidance. T.G.P.G. supervised the study and data analysis. All authors read and approved the final manuscript.

## SUPPLEMENTARY FIGURES AND TABLES

**Supplementary Figure 1:**
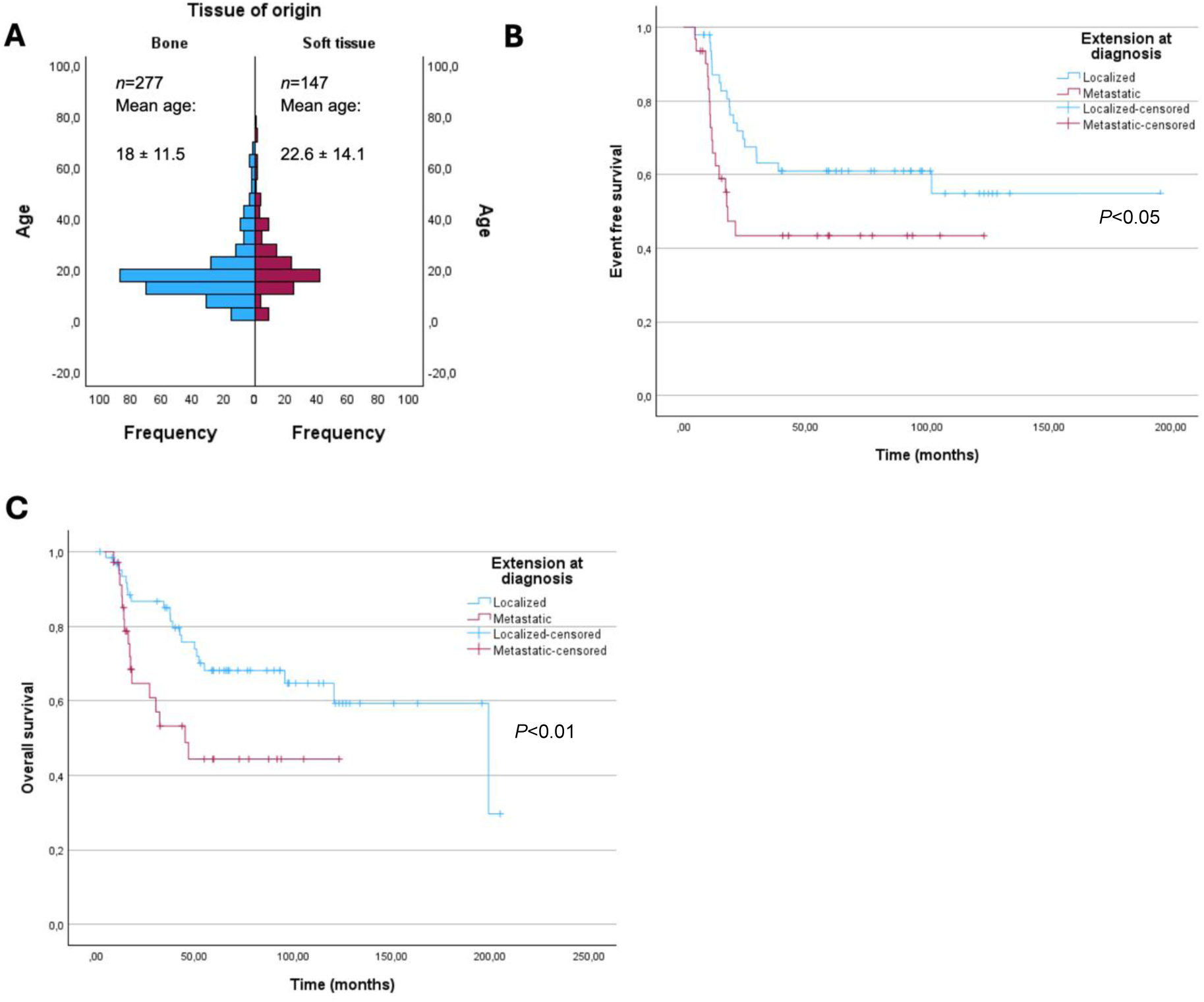
**A)** Comparison showing greater incidence of cases arising in soft tissue in older patients from the study cohort (Mann-Whitney U test *P*<0.001). **B and C)** Kaplan-Meier curves of EFS (61.4 ± 46.3 versus 33.7 ± 33.9 months; HR: 2.149; 95% CI: 1.16-3.98; *P*=0.015) and OS (72 ± 49.7 *versus* 37.6 ± 18.1 months; HR: 2.758; 95% CI: 1.46-5.2; *P*=0.002) showing poor prognosis association with metastatic presentation of the disease.

**Supplementary Figure 2:**
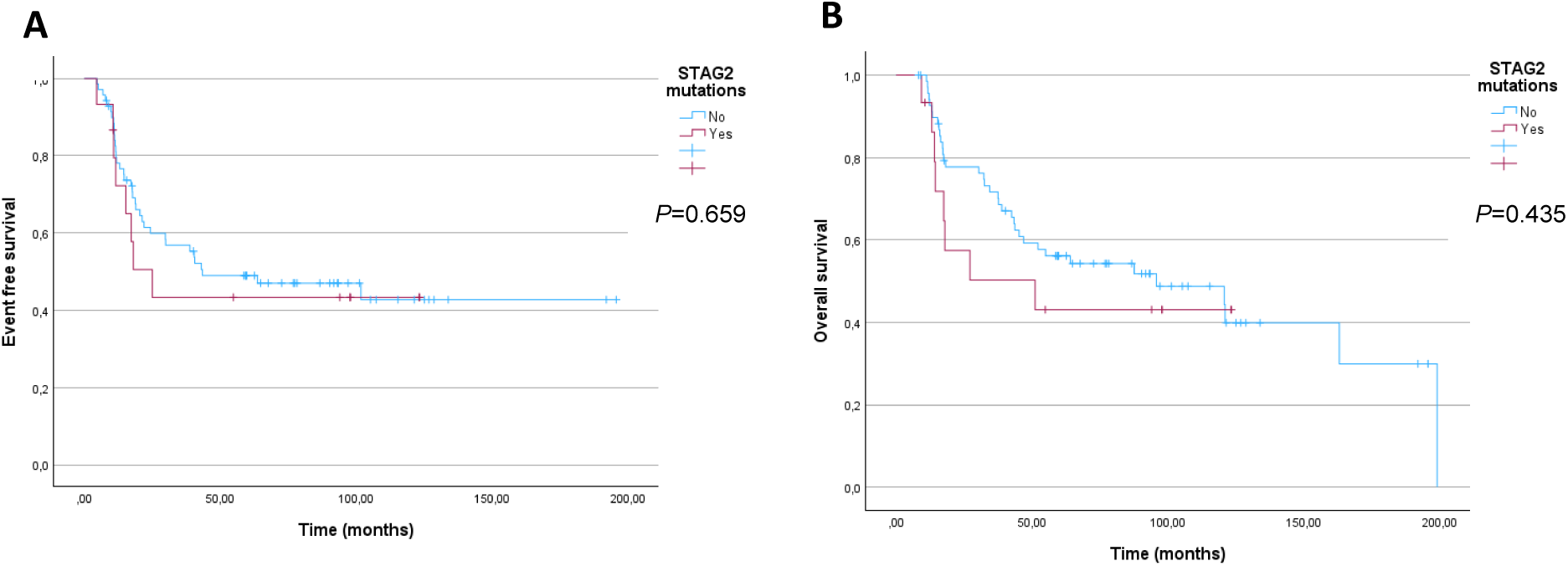
Kaplan-Meier analyses of differences in **(A)** event free survival (61.5±14.5 months in *STAG2* mutated *versus* 98.8±11 months in *STAG2* wt; HR 1.188; 95% CI:0.552–2.558; *P*=0.659) and **(B)** overall survival (64.9±13.8 months in *STAG2* mutated *versus* 104.9±10.7 months in *STAG2* wt; HR 1.331; 95% CI: 0.615–2.878; *P*=0.468) between *STAG2* mutated and *STAG2*wt EwS.

**Supplementary Figure 3:**
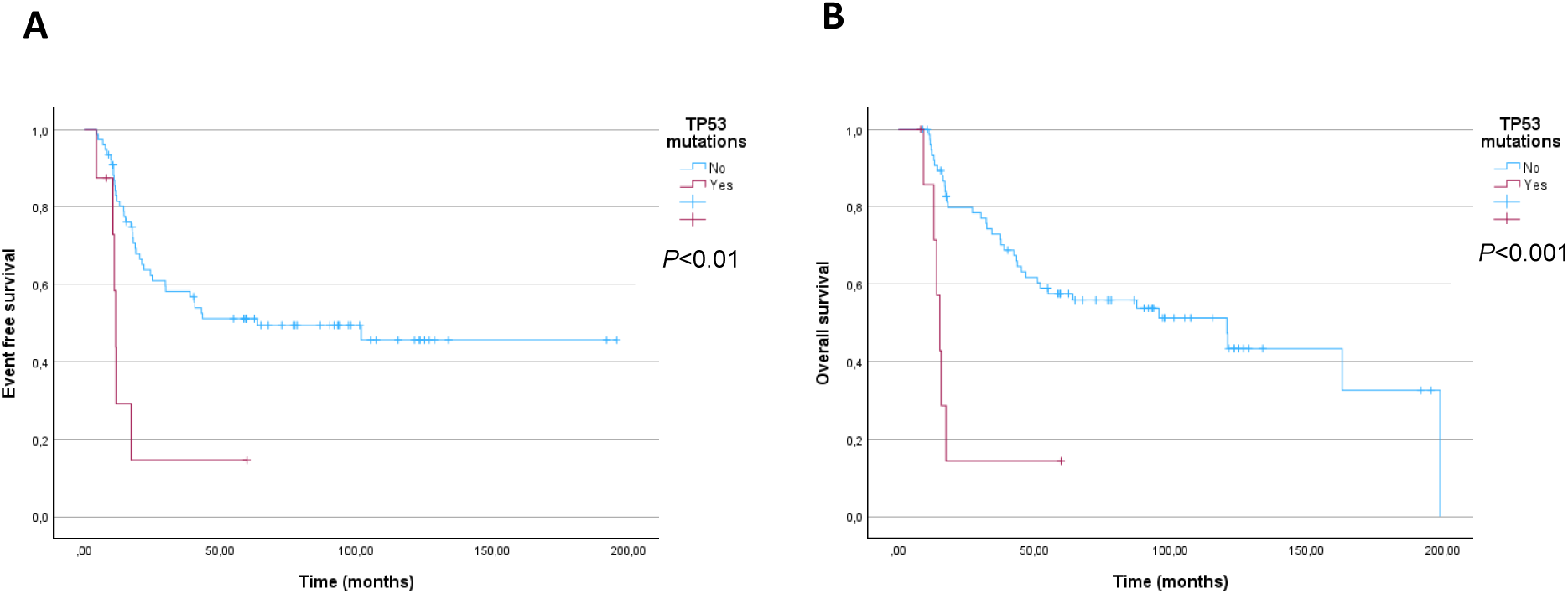
Kaplan-Meier representation of differences in **(A)** event free survival (18.4±6.6 months compared to 103.6±10.5 months for those *TP53*wt cases; HR 3.626; 95% CI:1.5–8.77; *P*<0.01) and **(B)** overall survival (20.7±6.1months *versus.* 109.3±10.25 months; HR 5.012; 95% CI:2.04–12.32; *P*<0.001) between *TP53* mutated and *TP53*wt EwS.

**Supplementary Figure 4:**
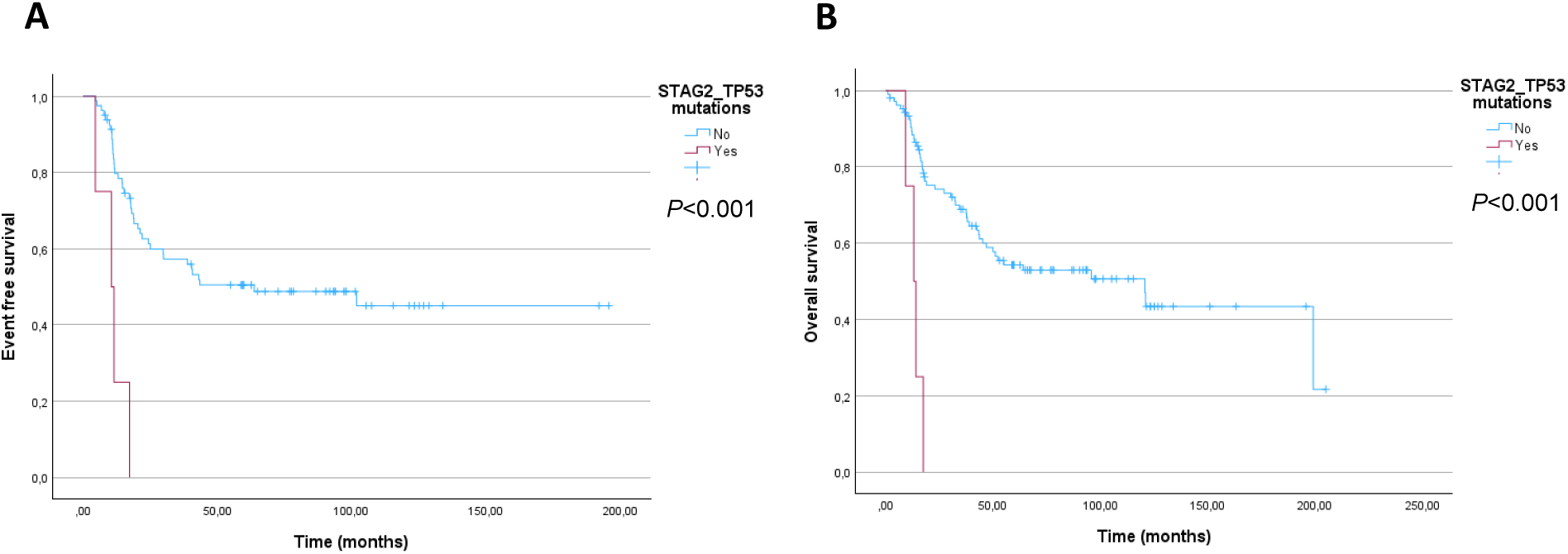
Kaplan-Meier representation of differences in **(A)** event free survival (11±2.6 months *versus* 102.3±10.3 months; HR 6.626; 95% CI: 2.25–19.47; *P*<0.001) and **(B)** overall survival (13.5±1.7 months *versus* 111± 9.4 months; HR 7.05; 95% CI: 2.39–20.75; *P*<0.001) between *STAG2*-*TP53* mutated and *STAG2*-*TP53*wt EwS.

**Supplementary Table 1.** Clinicopathological characteristics of EwS cases harboring EWSR1::FLI1 or EWSR1::ERG fusions.

| Clinical phenotypes and outcomes |  | Fusion type |  | P value |
| --- | --- | --- | --- | --- |
|  |  | EWSR1::FLI1 | EWSR1::ERG |  |
| Mean age at diagnosis (y.o.) |  | 20.5±12.6 | 22.4±13.6 | 0.42 |
| Sex | Male | 64.7% (205/317) | 63.4% (26/41) | 0.76 |
|  | Female | 35.3% (112/317) | 36.6% (15/41) |  |
| Tissue of origin | Bone | 62.2% (191/307) | 65.8% (27/41) | 0.76 |
|  | Soft tissue | 37.8% (116/307) | 34.1% (14/41) |  |
| Extension at diagnosis | Localized | 66% (208/315) | 79.5% (31/39) | 0.16 |
|  | Metastatic | 33.9% (107/315) | 20.5% (8/39) |  |
| Status | Alive | 50.4% (128/254) | 48.6% (18/37) | 0.77 |
|  | Dead of the disease | 48.4% (123/254) | 51.3% (19/37) |  |
|  | Dead of other causes | 1.2% (3/254) | 0 |  |

**Supplementary Table 2.** Coexistence of the main CNV observed in the study cohort (*n*=242). Numbers indicate the absolute count of cases with concomitant CNV.

|  | Gain_chr8 | Gain_chr12 | Gain_chr20 | Gain_chr1q | Loss_chr16q | Gain_chr2 | Gain_chr14 |
| --- | --- | --- | --- | --- | --- | --- | --- |
| Gain_chr8 |  | 52 | 40 | 38 | 32 | 37 | 32 |
| Gain_chr12 | 52 |  | 30 | 28 | 19 | 36 | 28 |
| Gain_chr20 | 40 | 30 |  | 23 | 17 | 24 | 22 |
| Gain_chr1q | 38 | 28 | 23 |  | 25 | 24 | 16 |
| Loss_chr16q | 32 | 19 | 17 | 25 |  | 14 | 22 |
| Gain_chr2 | 37 | 36 | 24 | 24 | 14 |  | 25 |
| Gain_chr14 | 32 | 28 | 22 | 16 | 16 | 25 |  |

## REFERENCES

1. Grünewald, T. G. P. et al. Ewing sarcoma. Nat. Rev. Dis. Primer 4, 5 (2018).

2. Mackintosh, C. et al. 1q gain and CDT2 overexpression underlie an aggressive and highly proliferative form of Ewing sarcoma. Oncogene 31, 1287–1298 (2012).

3. Dermawan, J. K. et al. Chromoplexy is a frequent early clonal event in EWSR1-rearranged round cell sarcomas that can be detected using clinically validated targeted sequencing panels. Cancer Res. https://doi.org/10.1158/0008-5472.CAN-23-2573 (2024) doi:10.1158/0008-5472.CAN-23-2573.

4. Nacev, B. A. et al. Clinical sequencing of soft tissue and bone sarcomas delineates diverse genomic landscapes and potential therapeutic targets. Nat. Commun. 13, 3405 (2022).

5. Tirode, F. et al. Genomic landscape of Ewing sarcoma defines an aggressive subtype with co-association of STAG2 and TP53 mutations. Cancer Discov. 4, 1342–1353 (2014).

6. Crompton, B. D. et al. The genomic landscape of pediatric Ewing sarcoma. Cancer Discov. 4, 1326–1341 (2014).

7. Brohl, A. S. et al. The genomic landscape of the Ewing Sarcoma family of tumors reveals recurrent STAG2 mutation. PLoS Genet. 10, e1004475 (2014).

8. Agelopoulos, K. et al. Deep Sequencing in Conjunction with Expression and Functional Analyses Reveals Activation of FGFR1 in Ewing Sarcoma. Clin. Cancer Res. Off. J. Am. Assoc. Cancer Res. 21, 4935–4946 (2015).

9. Zhang, N. et al. Molecular Heterogeneity of Ewing Sarcoma as Detected by Ion Torrent Sequencing. PloS One 11, e0153546 (2016).

10. Gillani, R. et al. Molecular Characterization Informs Prognosis in Patients With Localized Ewing Sarcoma: A Report From the Children’s Oncology Group. J. Clin. Oncol. 43, 3750–3759 (2025).

11. Chang, H.-Y. et al. Spectrum and Clinical Impact of Secondary Genetic Alterations in Translocation-Associated Sarcomas. *JCO Precis*. Oncol. 9, e2500603 (2025).

12. Soupir, A. et al. Genomic, transcriptomic, and immunogenomic landscape of over 1300 sarcomas of diverse histology subtypes. Nat. Commun. 16, 4206 (2025).

13. Surdez, D. et al. STAG2 mutations alter CTCF-anchored loop extrusion, reduce cis-regulatory interactions and EWSR1-FLI1 activity in Ewing sarcoma. Cancer Cell 39, 810–826.e9 (2021).

14. Adane, B. et al. STAG2 loss rewires oncogenic and developmental programs to promote metastasis in Ewing sarcoma. Cancer Cell 39, 827–844.e10 (2021).

15. Cancer Genome Atlas Research Network. Electronic address: & Cancer Genome Atlas Research Network. Comprehensive and Integrated Genomic Characterization of Adult Soft Tissue Sarcomas. Cell 171, 950–965.e28 (2017).

16. Cosci, I., Del Fiore, P., Mocellin, S. & Ferlin, A. Gender Differences in Soft Tissue and Bone Sarcoma: A Narrative Review. Cancers 16, 201 (2024).

17. Rochefort, P. et al. A Retrospective Multicentric Study of Ewing Sarcoma Family of Tumors in Patients Older Than 50: Management and Outcome. Sci. Rep. 7, 17917 (2017).

18. Pieper, S. et al. Ewing’s Tumors over the Age of 40 – a Retrospective Analysis of 47 Patients Treated According to the International Clinical Trials EICESS 92 and EURO-E.W.I.N.G. 99. Onkologie 31, 657–663 (2008).

19. Applebaum, M. A. et al. Clinical features and outcomes in patients with extraskeletal Ewing sarcoma. Cancer 117, 3027–3032 (2011).

20. Salah, S. et al. Outcomes of extraskeletal vs. skeletal Ewing sarcoma patients treated with standard chemotherapy protocol. Clin. Transl. Oncol. Off. Publ. Fed. Span. Oncol. Soc. Natl. Cancer Inst. Mex. 22, 878–883 (2020).

21. Orth, M. F. et al. Systematic multi-omics cell line profiling uncovers principles of Ewing sarcoma fusion oncogene-mediated gene regulation. Cell Rep. 41, 111761 (2022).

22. Boone, M. A. et al. Identification of a Novel FUS/ETV4 Fusion and Comparative Analysis with Other Ewing Sarcoma Fusion Proteins. Mol. Cancer Res. MCR 19, 1795–1801 (2021).

23. Anderson, N. D. et al. Rearrangement bursts generate canonical gene fusions in bone and soft tissue tumors. Science 361, eaam8419 (2018).

24. Lawrence, M. S. et al. Mutational heterogeneity in cancer and the search for new cancer-associated genes. Nature 499, 214–218 (2013).

25. Gillani, R. et al. Germline predisposition to pediatric Ewing sarcoma is uniquely characterized by inherited pathogenic variants in DNA damage repair genes. 2022.01.07.22268685 Preprint at 10.1101/2022.01.07.22268685 (2022).

26. Rao, R. C. & Dou, Y. Hijacked in cancer: the KMT2 (MLL) family of methyltransferases. Nat. Rev. Cancer 15, 334–346 (2015).

27. Casula, M. et al. Germline and somatic mutations in patients with multiple primary melanomas: a next generation sequencing study. BMC Cancer 19, 772 (2019).

28. Zhang, L. et al. Genomic Analysis of Nasopharyngeal Carcinoma Reveals TME-Based Subtypes. Mol. Cancer Res. MCR 15, 1722–1732 (2017).

29. Morin, R. D. et al. Genetic Landscapes of Relapsed and Refractory Diffuse Large B-Cell Lymphomas. Clin. Cancer Res. Off. J. Am. Assoc. Cancer Res. 22, 2290–2300 (2016).

30. Kohsaka, S. et al. A recurrent neomorphic mutation in MYOD1 defines a clinically aggressive subset of embryonal rhabdomyosarcoma associated with PI3K-AKT pathway mutations. Nat. Genet. 46, 595–600 (2014).

31. Ogura, K. et al. Prospective Clinical Genomic Profiling of Ewing Sarcoma: ERF and FGFR1 Mutations as Recurrent Secondary Alterations of Potential Biologic and Therapeutic Relevance. *JCO Precis*. Oncol. e2200048 (2022) doi:10.1200/PO.22.00048.

32. Sobczuk, P., et al. 47MO Activating EZH2 mutations define a new subset of aggressive Ewing sarcomas. ESMO Open 8, (2023).

33. López-Guerrero, J. A. et al. Clinicopathological significance of cell cycle regulation markers in a large series of genetically confirmed Ewing’s sarcoma family of tumors. Int. J. Cancer 128, 1139–1150 (2011).

34. Huang, H.-Y. et al. Ewing sarcomas with p53 mutation or p16/p14ARF homozygous deletion: a highly lethal subset associated with poor chemoresponse. J. Clin. Oncol. Off. J. Am. Soc. Clin. Oncol. 23, 548–558 (2005).

35. Casey, D. L. et al. TP53 mutations increase radioresistance in rhabdomyosarcoma and Ewing sarcoma. Br. J. Cancer 125, 576–581 (2021).

36. Paragji, A. et al. Revisiting CDKN2A dysregulation in Ewing sarcoma. Mol. Oncol. 19, 994–1001 (2025).

37. Zarrei, M., MacDonald, J. R., Merico, D. & Scherer, S. W. A copy number variation map of the human genome. Nat. Rev. Genet. 16, 172–183 (2015).

38. Maurici, D. et al. Frequency and implications of chromosome 8 and 12 gains in Ewing sarcoma. Cancer Genet. Cytogenet. 100, 106–110 (1998).

39. Hattinger, C. M. et al. Prognostic impact of chromosomal aberrations in Ewing tumours. Br. J. Cancer 86, 1763–1769 (2002).

40. Díaz-Martín, J. et al. Prospective evaluation of copy number alterations validates chromosome 1q gain as an independent marker of poor prognosis in localized Ewing sarcoma. Exp. Mol. Pathol. 144, 105008 (2025).

41. Ranft, A. et al. Potential biomarkers of Ewing sarcoma identified through a Europe-wide analysis of prospectively collected samples. Sci. Rep. 16, 11613 (2026).

42. Machado, I. et al. Sarcomas with EWSR1::Non-ETS Fusion (EWSR1::NFATC2 and EWSR1::PATZ1). Surg. Pathol. Clin. 17, 31–55 (2024).

43. Koelsche, C. et al. DNA methylation profiling distinguishes Ewing-like sarcoma with EWSR1-NFATc2 fusion from Ewing sarcoma. J. Cancer Res. Clin. Oncol. 145, 1273–1281 (2019).

44. Cheng, D. T. et al. Comprehensive detection of germline variants by MSK-IMPACT, a clinical diagnostic platform for solid tumor molecular oncology and concurrent cancer predisposition testing. BMC Med. Genomics 10, 33 (2017).

45. Pieper, S. et al. Ewing’s tumors over the age of 40: a retrospective analysis of 47 patients treated according to the International Clinical Trials EICESS 92 and EURO-E.W.I.N.G. 99. Onkologie 31, 657–663 (2008).

46. Bacci, G. et al. Adjuvant and neoadjuvant chemotherapy for Ewing sarcoma family tumors in patients aged between 40 and 60: report of 35 cases and comparison of results with 586 younger patients treated with the same protocols in the same years. Cancer 109, 780–786 (2007).

47. Worch, J. et al. Age dependency of primary tumor sites and metastases in patients with Ewing sarcoma. Pediatr. Blood Cancer 65, e27251 (2018).

48. Richardson, R. B. p53 mutations associated with aging-related rise in cancer incidence rates. Cell Cycle Georget. Tex 12, 2468–2478 (2013).

49. Shahzad, M. et al. What have we learned about TP53-mutated acute myeloid leukemia? Blood Cancer J. 14, 202 (2024).

50. Yanada, M. et al. TP53 mutations in older adults with acute myeloid leukemia. Int. J. Hematol. 103, 429–435 (2016).

51. Rücker, F. G. et al. TP53 alterations in acute myeloid leukemia with complex karyotype correlate with specific copy number alterations, monosomal karyotype, and dismal outcome. Blood 119, 2114–2121 (2012).

52. Bourgi, A., Vincentelli, A., Rusch, E. & Bruyère, F. Youth matters: a systematic review of the molecular and clinical landscape of bladder cancer in young adults. World J. Urol. 43, 321 (2025).

53. Duda-Madej, A., Lipska, P., Viscardi, S., Bazan, H. & Sobieraj, J. Targeting Skin Neoplasms: A Review of Berberine’s Anticancer Properties. Cells 14, 1041 (2025).

54. Liu, K. X. et al. Risk stratification by somatic mutation burden in Ewing sarcoma. Cancer 125, 1357–1364 (2019).

55. Wei, G. et al. Prognostic impact of INK4A deletion in Ewing sarcoma. Cancer 89, 793–799 (2000).

56. Lerman, D. M. et al. Tumoral TP53 and/or CDKN2A alterations are not reliable prognostic biomarkers in patients with localized Ewing sarcoma: a report from the Children’s Oncology Group. Pediatr. Blood Cancer 62, 759–765 (2015).

57. Ozaki, T. et al. Genetic imbalances revealed by comparative genomic hybridization in Ewing tumors. Genes. Chromosomes Cancer 32, 164–171 (2001).

58. Walker, V. et al. Gene partners of the EWSR1 fusion may represent molecularly distinct entities. Transl. Oncol. 38, 101795 (2023).

59. Bose, R. et al. ERF mutations reveal a balance of ETS factors controlling prostate oncogenesis. Nature 546, 671–675 (2017).

60. Richter, G. H. S. et al. EZH2 is a mediator of EWS/FLI1 driven tumor growth and metastasis blocking endothelial and neuro-ectodermal differentiation. Proc. Natl. Acad. Sci. 106, 5324–5329 (2009).

